# The Experiences of Sexual Minority Service Users and Staff in Statutory Mental Health Services in the United Kingdom: a Systematic Review and Narrative Synthesis

**DOI:** 10.1101/2025.09.05.25335187

**Authors:** Liam Mackay, Chia-yi Lin, Merle Schlief, David Osborn, Helen Killaspy

## Abstract

**Background:** Despite improving legal protections in recent decades for sexual minorities in the UK, sexual minorities continue to experience significant mental health disparities relative to heterosexuals, and report negative experiences within mental health services that is related to their sexual identity. However, little attention has been paid to the impact of these negative experiences on outcomes within services, or to the experiences of sexual minority staff. To overcome these gaps, the aim of this review is to explore the experiences of sexual minority staff and service users in statutory mental health services in the UK, including a focus on outcomes for service users.

**Methods:** For the systematic search, we searched: Applied Social Sciences Index & Abstracts, EMBASE, MEDLINE®, PsycINFO, CINAHL Plus, and Web of Science for relevant articles, and for the suppementaty search, we searched the websites of relevant organisations, in addition to searching Google, Google Scholar, and DuckDuckGo. We included all study designs, published from 2010 onwards, that included first-hand accounts of sexual minority staff or service users in statutory mental health services in the UK. We used a narrative synthesis to synthesise the findings.

**Results:** Of 7427 articles identified, 84 were assessed at full text screening, with 11 being included in the review. 9 of the included studies were with adults, with the majority being conducted in the context of Talking Therapies and primary care counselling services (n = 7). Across studies, sexual minorities reported anticipatory anxiety about experiencing prejudice and discrimination in services, microaggressions (e.g., heteronormative assumptions) and the pathologization of their sexual minority identities. There are mixed findings about whether certain sexual minority groups have worse outcomes in Talking Therapies services. We found no studies that explored the experiences of sexual minority staff.

**Conclusions:** Sexual minority service users are reporting negative experiences in some statutory mental health services in the UK that are related to their sexual identity. In line with service user recommendations, services should consider introducing sexual minority training for staff to improve the experiences of service users. An absence of research on sexual minority staff, and little research on the experiences of young people from sexual minorities, highlight the need for research in this area

## Introduction

In the UK, mental health services have been developed against the historical backdrop of homosexuality being viewed as a pathological condition. This view of homosexuality as an illness legitimised ‘treatments’ that attempted to change homosexuals into heterosexuals (King & Bartlett, 1999). Despite the partial decriminalisation of homosexuality in England in Wales under the Sexual Offences Act of 1967, treatments that attempted to change the sexual orientation of same sex attracted people reached a peak in the 1960s and 70s in the UK (Smith et al., 2004). This is in line with the diagnostic nomenclature, with homosexuality being removed as a diagnosis from the DSM in 1973, and finally from the ICD in 1990 (Drescher, 2015).

In the decades since, there has been significant legal progress for sexual minorities (e.g., lesbian, gay, bisexual, queer, pansexual, asexual; LGBQ+) in the UK, including the legalisation of same-sex marriage, and improving societal attitudes (National Centre for Social Research, 2023). Legislation has also been introduced to protect sexual minorities from discrimination, victimisation, and harassment in different settings, including in public services such as (mental) health services (The Equality Act 2010). Despite this, evidence from population cohort studies has shown that sexual minorities experience around double the rate of common mental disorders (CMDs), such as depression and anxiety, as well as a higher rate of self-harm and substance misuse (Amos et al., 2020; Kidd et al., 2024; King et al., 2008; Pitman et al., 2021; Plöderl & Tremblay, 2015; Semlyen et al., 2016). There is evidence that this mental health disparity emerges as young as aged 10, with young people who grow up to identify as a sexual minority having significantly higher depression symptoms at aged 10 relative to those who grow up to identify as heterosexual (Irish et al., 2019). Young people from sexual minorities also have increased risk of psychotic like experiences from age 12, and over 4.5 times the odds of self-harm with suicidal intent at age 21, relative to heterosexuals (Corcoran et al., 2024; Irish et al., 2019). Research suggests that these inequalities are worse for older sexual minority adults, and for bisexuals (Pitman et al., 2021; Semlyen et al., 2016).

The dominant framework for understanding this mental health disparity is Minority Stress Theory (Brooks, 1981; Meyer, 2003). This suggests that sexual minorities experience unique and chronic stressors as a result of living in stigmatizing social environments. These stressors are suggested to exist on a continuum from distal stressors within the social environment, including victimisation and discrimination, to proximal stressors that are internal, such as internalised homophobia or identity concealment. It is exposure to these unique chronic stressors, in addition to general stressors experienced by everyone regardless of minority status, that is believed to contribute to the mental health inequalities between sexual minorities and heterosexuals (Frost & Meyer, 2023).

In light of the increased psychological morbidity experienced by sexual minorities, there has been research to investigate whether sexual minorities utilise mental health services at a higher rate than heterosexuals. An evidence review commissioned by the UK government found that sexual minorities were more likely to use mental health service compared to heterosexuals (Hudson & Metcalf, 2016). However, despite sexual minorities potentially higher rates of mental health services use, there is evidence that services are not always meeting the needs of sexual minority service users, with some sexual minorities reporting negative experiences within services that is related to their sexuality.

Systematic reviews of the international literature have found that sexual minorities report both general negative experiences of services, such as costly private services and difficult to access public services, in addition to negative experiences relating to their identity (McNamara & Wilson, 2020; River et al., 2025). These experiences include people from sexual minorities reporting that their sexual identity was pathologized by practitioners, experiences of heteronormative assumptions (i.e., the assumption that people are heterosexual), and being faced with practitioners who lacked knowledge about sexual identities.

Whilst these reviews highlight that people from sexual minorities report negative experiences within mental health services that it is related to their identity, their focus was on experiences rather than outcomes of accessing mental health care. This means that it is unclear the impact, if any, that these negative experiences have on outcomes such as satisfaction with services and treatment outcomes. It was also out with the scope of these reviews to explore the experiences of sexual minority staff.

Results from the UK’s National Health Service (NHS) staff survey highlight the importance of considering sexual minority staff, with the results of the survey showing that sexual (and gender) minority staff reported disproportionately poorer experiences of working in the NHS, and were more likely report experiences of bullying, violence, and discrimination relative to heterosexual staff (NHS Employers, 2024). These findings are consistent with those of a survey conducted by the Royal College of Psychiatrists, the professional body in the UK that holds responsibility for representing and training psychiatrists, which found that sexual (and gender) minority psychiatrists reported experiences of bullying and harassment in the workplace (Royal College of Psychiatrists, 2022).

To address the lack of attention in previous reviews to the experiences of sexual minority staff and outcomes for sexual minorities within mental health services, this review aimed to explore the experiences of sexual minority staff and service users in statutory mental health services in the UK. We were interested in qualitative, quantitative, and mixed methods research that had explored people’s experiences of accessing services, in addition to their satisfaction with services and treatment outcomes. As people from sexual minorities are more likely to experience common mental disorders (i.e., anxiety and depression) relative to heterosexuals, we included research conducted within psychological services and primary care counselling services. We were also interested in the experiences of sexual minority staff working within statutory mental health services.

In line with these aims, our review questions were:

1. What are the experiences of sexual minority patients accessing statutory mental health services in the UK in relation to their sexual minority status?
2. What are the experiences of sexual minority staff who work in statutory mental health services in the UK in relation to their sexual minority status?

## Methods

The protocol for this review was registered on PROSPERO (CRD42024570682) and results are reported in line with the Preferred Reporting Items for Systematic Reviews and Meta-Analyses (PRISMA) checklist (Page et al., 2021).

### Search Strategy

We created the search terms and developed the search strategy in consultation with a subject specialist librarian at out institution. The following databases were searched: Applied Social Sciences Index & Abstracts (ProQuest), EMBASE (Ovid), MEDLINE® (Ovid), PsycINFO (Ovid), CINAHL Plus (EBSCO Host), and Web of Science (Core Collection). No language restrictions were imposed, but we only included papers from 2010 onwards, in line with the introduction of The Equality Act (2010), due to the legal requirement that this current legislation places on public services to eliminate discrimination for sexual minorities (and those with other protected characteristics under the act).

Following the initial development of the search terms, the primary author piloted the search strategy to ensure that it captured papers that we were aware of that met our inclusion criteria. Further iterations were developed by adding more search terms, in consultation with the subject specialist librarian, before piloting the strategy again.

The search terms were developed around three concepts:

**Concept 1:** Sexual minorities (e.g., lesbian, gay, bisexual, queer).

**Concept 2:** Experiences of services. In this concept we captured experiences of receiving support from statutory mental health services (e.g., experience* of service user*).

**Concept 3:** Statutory mental health services. This included psychological therapy delivered within primary care, and within NHS Talking Therapies for anxiety and depression services (services for adults in England experiencing CMDs), in addition to secondary, community, and inpatient mental health services.

A combination of key words (Appendix I), and MeSH terms/Subject Headings were used. The initial search was conducted on 29th July 2024. This was rerun on the 18^th of^ July 2025, before submitting the review for publication, to capture any new papers published since the initial search. The results of the database searches were exported to Zotero, and then exported to Systematic Review Accelerator (an automated tool) for deduplication using the relaxed filter (Clark et al., 2020). This filter was chosen as it does not mislabel non-duplicates at the cost of missing a small number of duplicates and is recommended for reviews with over 2000 results. The relaxed filter does not require the results to be manually checked, however, to be cautious, the primary author (LM) manually checked the results to ensure that the tool had correctly identified duplicates and hadn’t mislabelled non-duplicates as duplicates. The deduplicated results were then exported to the systematic review management software, Covidence.

The following websites were also searched for any relevant grey literature publications: NHS Confederation, Stonewall, Royal College of Psychiatrists, Royal College of Nursing, British Psychological Society, and the LGBT Foundation. Grey literature searches were also conducted on the search engines Google, Google scholar, and DuckDuckGo, using combinations of search terms capturing the three concepts (e.g., lgb* AND service experience* AND mental health service*). The first 100 results were searched. Forwards and backwards citation searching was also conducted by scanning the reference lists of included studies, and by searching on Web of Science (Core Collection) for any included studies to find any relevant papers that had cited the included study. In situations where the full text of peer reviewed publications or grey literature could not be retrieved, or further information was required to assess eligibility, the primary author (LM) contacted the corresponding author to request access.

### Study Selection and Eligibility Criteria

We included papers reporting on studies of any design, but excluded systematic reviews, position statements, books, conference abstracts, editorials, commentaries, and other opinion pieces. We were interested in studies that explored the experiences of sexual minority staff and users of statutory mental health services in the UK. Statutory mental health services are predominantly provided by the NHS in the UK, and are delivered in primary care settings (e.g., GP practices), community settings (e.g., community mental health teams), outpatient community centres or hospital settings (e.g., adult mental health teams), specialist services (e.g., early intervention for psychosis services), and inpatient settings. We also included psychological therapy delivered in primary care counselling services, and within Talking Therapies services for anxiety and depression (formerly called Improving Access to Psychological Therapies; IAPT). Some private and not-for-profit organisations are also commissioned by the NHS to provide mental health services and studies conducted in these settings were potentially eligible for inclusion.

Sexual minority status could be reported in various ways including by identity (e.g., lesbian, gay, bisexual, queer), behaviour (e.g., men who have sex with men), and/or attractions (e.g., women who are attracted to women). We included studies that included heterosexual individuals and those with a minority gender identity (e.g., transgender and non-binary) if the results for those with a minority sexual orientation were reported separately. Studies were excluded if they only focussed on gender minorities without mentioning sexual orientation.

We took a broad definition of ‘experiences of services’ and included any studies that explored experiences in relation to individuals’ sexual minority status. For service users, experiences of services could include treatment outcomes, experiences of accessing and receiving services (including experiences of prejudice and discrimination) and satisfaction with services. For staff, studies could investigate any theme relevant to their sexual minority status, such as satisfaction with working within services, or experiences of discrimination or prejudice in the workplace. We only included studies that explored the first-hand accounts of sexual minority service users and staff.

Following the exportation of the deduplicated results to Covidence, the primary author conducted title and abstract screening, with a second reviewer (CL) screening 25% of results at this stage. Conflicts were resolved through discussion, in consultation with a third researcher (MS). LM and CL then independently conducted full text screening of all studies retained after title and abstract screening. Any disagreements about inclusion at this stage were resolved through discussion with two senior co-authors (HK & DO) to reach consensus agreement.

### Data Extraction and Quality Assessment

Data extraction was carried out using an Excel extraction template created by LM. Two researchers (LM & CL) piloted the template by extracting the data for one study, comparing these extractions, and amending the template accordingly. Data were then extracted for all included studies. This included the author(s), the year the study was published, title, study aim(s), study design, study methods (sample recruitment, outcomes and outcome measures), sample size, sexual orientation of participants, age of participants, mental health service setting, and a summary of the results.

Each included study was assessed for quality using the QualSyst tool (Kmet et al., 2004). In a slight deviation from the protocol, where we planned to assess included mixed methods studies with the Mixed Methods Appraisal Tool (MMAT; Hong et al., 2018), we also used the QualSyst tool to assess the quality of the included mixed methods study. This is because only one mixed methods study was included, and the MMAT does not allow for a summary score to be calculated meaning it was challenging to compare the quality of the mixed methods study with the other included studies.

Different versions of the QualSyst tool can be used to assess qualitative studies or quantitative studies with each item rated ‘yes’, ‘no’, or ‘partial’. An overall summary score is calculated for each study by summing the number of ‘yes’ responses (which receive 2 points), the number of ‘partial’ responses (which receive 1 point), and the number of ‘no’ responses (which receive 0 points), and then dividing this by the maximum possible score in order to calculate a score out of 100 for each study. For the mixed methods study, we used the quantitative and qualitative checklists for the respective components of the study to calculate a score for each component. The quality assessment was completed independently by two researchers (LM & CL) for each study.

### Data Synthesis

We used a narrative synthesis to synthesise the findings, following elements of the general framework for narrative synthesis by Popay et al. (2006). This included developing a preliminary synthesis, exploring relationships in the data, and assessing the robustness of the synthesis. Using tools and techniques outlined by Popay et al. (2006), LM conducted the preliminary synthesis by tabulating the findings from each study in order to describe the findings and begin identifying patterns across the different studies. This preliminary synthesis was checked by the research team. Relationships in the data were explored by comparing similarities and differences across different studies and exploring whether results differed according to different contexts and/or populations. Finally, following Popay et al.’s (2006) guidance, we reflected critically on the synthesis process in order to assess the robustness of the synthesis. This involved considering: the method of synthesis used and its limitations, the quality, validity, and generalisability of the included studies, and any discrepancies or uncertainties identified.

## Results

Figure 1 shows the PRISMA flow diagram for this review. A total of 7427 studies were found in the systematic search, of which 3160 were duplicates. After screening 4267 at title and abstract screening, 84 were assessed at full text screening, with 8 being included in the review. An additional 3 studies were included following citation searching as part of the supplementary search, bringing the total number of studies included in the review to 11. Table 1 shows the characteristics of the included studies.

**Figure 1.**
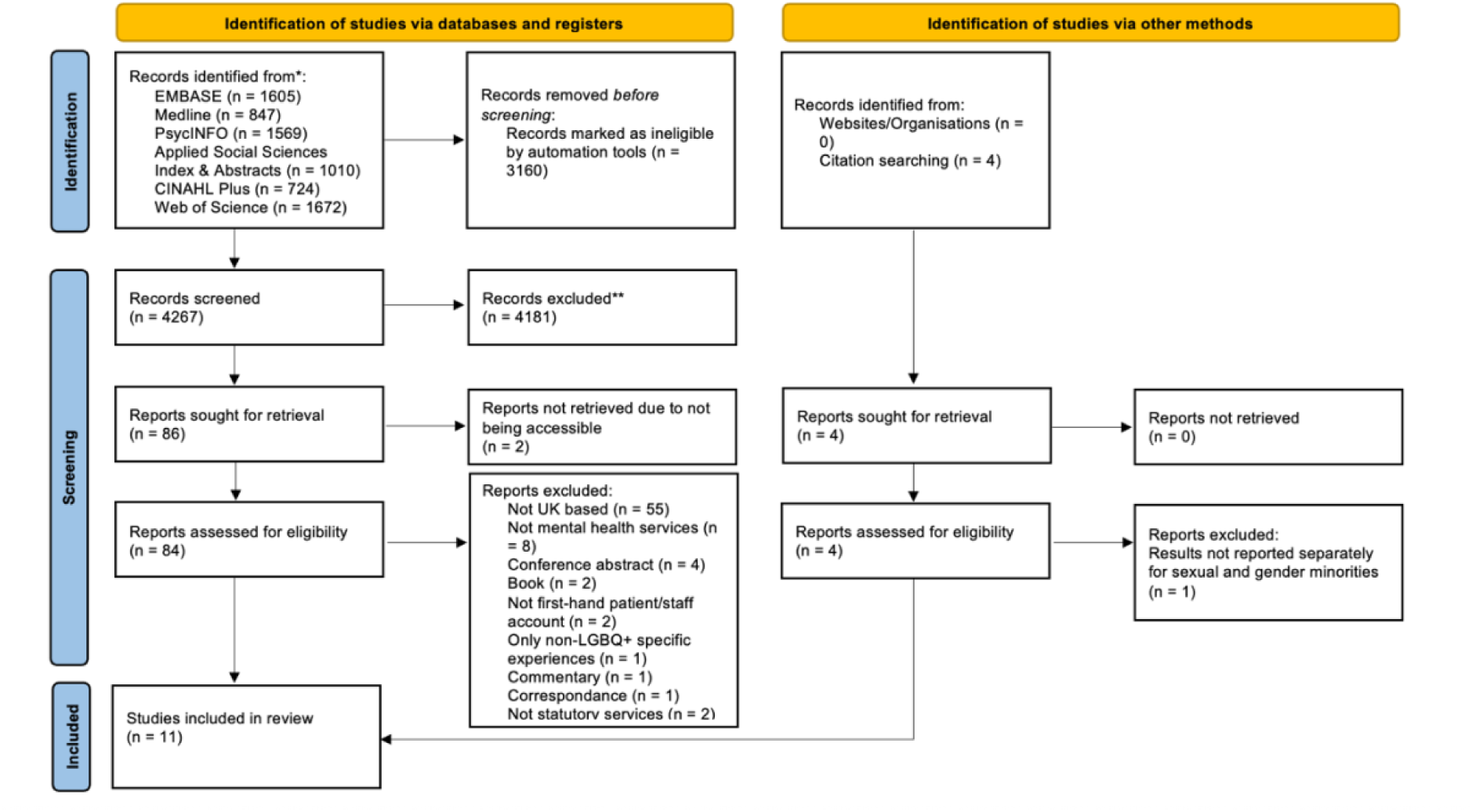
PRISMA Flow Diagram

**Table 1.** Characteristics of Included Studies.

| Author(s) | Aim(s) of study | Study design | Sample size, sexual orientation of participants | Age of participants, age range, M (SD) | Setting | Quality Assessment |
| --- | --- | --- | --- | --- | --- | --- |
| Foy et al., 2019 | To explore LGBTQ+ adults experiences of accessing and receiving psychological interventions from IAPT or primary care counselling, and their recommendations on how to improve services for sexual minorities | Mixed methods – used an online questionnaire with fixed-choice and optional open-ended questions. Fixed-choice responses were summarised using descriptive statistics, and open-ended responses were analysed using thematic analysis. | n = 8<br>6 Bisexual (75%)<br>2 Pansexual (25%)<br>2 Queer (25%)<br>*Participants could select more than one sexual orientation | 29.6 (age range and SD not reported) | IAPT services in England, and primary care counselling services in the UK | Quantitative component: 83.3/100<br>Qualitative component: 80.0/100 |
| Rimes et al., 2018 | To compare sociodemographic, clinical characteristics, and treatment outcomes, between lesbian, gay, bisexual, and heterosexual individuals receiving a primary care psychological intervention for common mental health problems | Quantitative - prospective cohort study using routinely collected clinical data. | n = 10,791<br>7047 women, who identified as:<br>188 Lesbian (2.7%)<br>222 Bisexual (3.2%)<br>6637 Heterosexual (94.2%)<br>3744 men, who identified as:<br>645 Gay (17.2%)<br>75 Bisexual (2.0%)<br>3024 Heterosexual (80.8%) | For women - M age (SD):<br>Lesbian women - 35.5 (SD = 11.1)<br>Bisexual women - 31.3 (SD = 10.3)<br>Heterosexual women - 38.0 (SD = 12.6)<br>For men -<br>Gay men - 38.0 (SD = 9.8)<br>Bisexual men - 35.0 (SD = 12.6)<br>Heterosexual men - 39.3 (SD = 12.1) | IAPT services in four boroughs in South London (Southwark, Lambeth, Lewisham, and Croydon) provided by South London and Maudsley NHS Foundation Trust | 95.5/100 |
| Morris et al., 2022 | To investigate sexual minorities experiences with attempting to access, and receiving, psychological services through IAPT in England, or primary care counselling services throughout the UK, for mild to moderate psychological difficulties, and to explore service users' perspectives of | Qualitative – Data was collected using semi-structured interviews and was analysed using thematic analysis. | n = 26<br>:<br>7 Gay (26.9%); of whom 6 (85.7%) identified as male and 1 (14.3%) as a genderqueer man<br>9 Lesbian (34.6%), of whom all identified as women<br>6 Bisexual (23.1%), of whom 2 (33.3%) identified as male and 4 (66.7%) identified as female<br>3 Queer (11.5%), of whom all identified as female<br>1 Pansexual (3.8%), | 31.6 (19 - 70, SD not reported) | IAPT services in England, and primary care counselling services in the UK | 85.0/100 |
|  | how services can be developed to optimise experiences and outcomes for sexual minorities with psychological problems |  | who identified as male |  |  |  |
| Rimes et al., 2019 | To explore the effects of minority sexual orientation, the interaction between minority sexual orientation and sex, and the interaction between minority sexual orientation and racial minority status, on treatment outcomes within IAPT services in England | Quantitative - prospective cohort study using routinely collected clinical data | n = 132, 923<br>85,831 women:<br>83,482 Heterosexual (97.3%)<br>1285 Lesbian (1.5%)<br>1064 Bisexual (1.2%)<br>47,092 men:<br>44,969 Heterosexual (95.5%)<br>1734 Gay (3.7%)<br>389 Bisexual (0.8%) | For women - age range, M (SD):<br>Heterosexual: 16 - 90, M = 41.4 (SD = 15.3)<br>Lesbian women: 16 - 83, M = 34.5 (SD = 12.4)<br>Bisexual women: 16 - 73, M = 28.2 (SD = 10.6)<br>For men - age range, M (SD):<br>Heterosexual: 16 - 90, M = 42.5 (SD = 14.8)<br>Gay: 16 - 90, M = 36.9 (SD = 12.8)<br>Bisexual: 16 - 70, M = 32.9 (SD = 13.5) | IAPT services in England | 95.5/100 |
| Robertson et al., 2015 | To explore the intimate relationship experiences of LGB people whilst an inpatient on a NHS psychiatric ward, and explore | Qualitative – Data collected using semi-structured interviews and analysed using Interpretative Phenomenological Analysis. | n = 6<br>3 Gay men (50%)<br>3 Lesbian women (50%)<br>Participants had diagnoses that | 31 - 57, M = 45 (SD not reported) | Inpatient psychiatric wards in the UK provided by the NHS | 70.0/100 |
|  | how these experiences influence recovery |  | included schizophrenia, bipolar affective disorder, and personality disorder |  |  |  |
| Lloyd et al., 2021 | To explore LGBTQ+ adults experiences of a LGBTQ+ Wellbeing Group | Qualitative - data collected using an online survey and responses analysed using qualitative content analysis. | n = 18<br>11 participants provided their sexual orientation:<br>7 Gay (63.3% of those who provided their sexual orientation)<br>2 Lesbian (1.8%)<br>1 Unsure (0.1%)<br>1 Other (0.1%)<br>Gender of participants (n = 11):<br>7 Male (63.3%)<br>4 Female (36.6%) | M = 28.5 (11.8) | Talking Therapies Southwark (an IAPT service in South London) | 90.0/100 |
| Hambrook et al. 2022 | (1) To describe and compare the sociodemographic clinical characteristics of patients attending and dropping out of an LGBTQ+ wellbeing group; (2) To explore changes in outcome measures following the LGBTQ+ Wellbeing Group | Quantitative – Used routinely collected clinical data that was collected as part of an LGBTQ+ wellbeing group delivered in a Talking Therapies service in South London | n = 78<br>53 Gay men (67.9%)<br>13 Lesbian women (16.7%)<br>4 Bisexual men (5.1%)<br>2 Bisexual women (2.6%)<br>6 Uncertain/undisclosed (7.7%) | 21 - 59, M = 35.4 (SD = 10.8) | Talking Therapies Southwark (a Talking Therapies service in South London) | 81.8/100 |
| Crawford et al., 2016 | To determine the prevalence of and | Quantitative - Cross-sectional | n = 14, 587 | 14, 148 participants reported their age. | Primary and secondary | 77.3/100 |
|  | risk factors for perceived negative effects of psychological treatment for common mental disorders | survey. Data was collected as part of the second round of the National Audit of Psychological Therapies in England and Wales in 2012 - 2013. The audit includes an examination of routine clinical records and a survey of individuals using primary and secondary care services. This was supplemented with information from the national IAPT programme in England. | 13, 532 provided data on sexual orientation. These participants identified as:<br>12, 874 Heterosexual (95.1%)<br>365 Lesbian/Gay (2.7%)<br>320 Bisexual/Other (2.4%) | The Mean age and SD are not reported.<br>1088 (7.7%) aged 18 - 24<br>2513 (17.8%) aged 15 - 34<br>3287 (23.2%) aged 35 - 44<br>3519 (24.9%) aged 45 - 54<br>2474 (17.5%) aged 55 - 64<br>980 (6.9%) aged 65 - 74<br>287 (2.0%) aged 75+ | psychological treatment services in England and Wales |  |
| Camp et al., 2024 | To investigate treatment outcomes and completion for SGM adolescents and their cisgender and heterosexual peers, in the National & Specialist CAMHS, DBT Service | Quantitative - mixed independent and repeated measures design using routinely collected data | For individuals who dropped out at pre-treatment, there were:<br>10 bisexual/pansexual individuals<br>6 gay/lesbian individuals<br>6 heterosexual individuals<br>For the individuals who dropped out in during treatment, there were: | For individuals who dropped out at pre-treatment, the M (SD) by each sexual orientation was:<br>Bisexual/pansexual: 16.9 (SD = 0.94)<br>Gay/lesbian: 16.7 (SD = 1.09)<br>Heterosexual: 16.4 (SD = 0.8)<br>For individuals who dropped out during treatment: | A National & Specialist CAMHS DBT service provided by the NHS | 95.5/100 |
|  |  |  | 13<br>bisexual/pansexual individuals<br>2 gay/lesbian individuals<br>7 heterosexual individuals<br>For individuals who completed treatment, there were:<br>46<br>bisexual/pansexual individuals<br>15 gay/lesbian individuals<br>36 heterosexual individuals | Bisexual/pansexual: 16.5 (SD = 1.0)<br>Gay/lesbian: 14.2 (SD = 0.1)<br>Heterosexual: 16.3 (SD = 0.8)<br>For individuals who completed treatment:<br>Bisexual/pansexual: 16.5 (SD = 1.0)<br>Gay/lesbian: 16.2 (SD = 1.0)<br>Heterosexual: 16.3 (SD = 1.0) |  |  |
| Camp et al., 2023 | To understand adolescents' perceptions of how well DBT met their needs as a GSM individual and how effective the approach was for supporting them with GSM-associated difficulties, and suggestions for improvements in how DBT could better support GSM young people | Qualitative - Data was collected using semi-structured interviews and analysed using reflexive thematic analysis | 14 young people, who identified their sexual orientation as:<br>6 bisexual (42.9%)<br>3 Gay/lesbian (21.4%)<br>2 Pansexual (14.3%)<br>1 Queer (7.1%)<br>1 bisexual/asexual (7.1%)<br>1 aromantic/queer (7.1%) | 17.4 (SD = 1.03) | A National & Specialist CAMHS DBT service provided by the NHS | 90.0/100 |
| Kent et al., 2025 | To describe pretreatment characteristics and treatment outcomes for | Quantitative retrospective cohort study using routine clinical data | 100, 389 included in the analysis, of which 94, 239 had indicated their sexual | Bisexual men: 30.4 (SD = 10.7)<br>Gay men: 35.6 (SD = 11.7) | Seven Talking Therapies services in North and | 100/100 |
|  | depression and anxiety between LGB and heterosexual patients, stratified by gender, and to evaluate differences in recovery, improvement, engagement, and attrition rates between these two groups |  | orientation. These were reported as:<br>557 bisexual men (0.6%)<br>2725 gay men (2.9%)<br>26, 017 heterosexual men (27.6%)<br>2867 bisexual women (2.4%)<br>1273 lesbian women (1.4%)<br>60, 799 heterosexual women (64.5%)<br>1185 reported being unsure (1.2%)<br>4975 declined to answer when asked about their sexual orientation (5.0%) | Heterosexual men: 38.4 (SD = 13.6)<br>Bisexual women: 27.1 (SD = 7.1)<br>Lesbian women: 33.1 (SD = 11.6)<br>Heterosexual women: 37.1 (SD = 13.6) | Central East London |  |

*IAPT = Improving Access to Psychological Therapies (now called Talking Therapies for Anxiety and Depression)*
*LGB = Lesbian, Gay, and Bisexual*
*LGBQ+ = Lesbian, Gay, Bisexual, Queer +*
*GSM = Gender and Sexual Minorities*
*DBT = Dialectical Behaviour Therapy*

Of the included studies, 6 were quantitative, 4 were qualitative, and 1 was mixed methods. Nine of the included studies were with adults. Seven of these were conducted in Talking Therapies for anxiety and depression services (called Improving Access to Psychological Therapies services at the time of some of the included studies being conducted, but from this point onwards referred to as Talking Therapies services) and primary care counselling services (n = 7), one was conducted in inpatient mental health wards (n = 1), and one explored service users’ experiences of primary and secondary care services (n = 1). Two of the included studies explored the experiences of young people from sexual minorities within a national and specialist Dialectical Behaviour Therapy (DBT), Child and Adolescent Mental Health Service (CAMHS), provided by the NHS (n = 2). All of the included studies explored the experiences of sexual minority service users. We found no studies that explored the experiences of sexual minority staff in statutory mental health services in the UK.

The full quality assessment is reported in Appendices II and III. The included studies were generally of high quality, with all scoring at least 70 (out of 100) according to the quality assessment tool. Of the quantitative studies, the study by Crawford et al., (2016) was rated to be of lower quality (scoring 77.3) due to the subject characteristics not being sufficiently described, the participant recruitment strategy being likely to introduce bias, and the outcome measure not being well defined.

Three of the quantitative studies were of particularly good quality, scoring over 95 (Kent et al., 2025; Rimes et al., 2018, 2019). The qualitative study conducted in inpatient settings scored 70.0 as there was no evidence of verification procedures to establish the credibility of the study findings and no report on reflexivity (Robertson et al., 2015), whilst the other included qualitative studies all scored 85.0 or above (Camp et al., 2023; Lloyd et al., 2021; Morris et al., 2022; Pitt et al., 2024).

The quantitative component of the mixed methods study scored 83.3 and was scored down slightly due to the use of an unvalidated questionnaire, and the possibility of the sampling strategy introducing response bias. The qualitative component scored 80.0 and was partly scored down due to the lack of reflexivity statement (Foy et al., 2019).

The table in Appendix IV shows the preliminary synthesis which was conducted by tabulating the findings from each of the included studies (Popay et al., 2006).

We first report the studies exploring the experiences of adults from sexual minorities. Two of the qualitative studies, and the mixed methods study, were particularly rich sources regarding the experiences of adults from sexual minorities in statutory mental health services. These were conducted in primary care counselling services and Talking Therapies services (Foy et al., 2019; Morris et al., 2022) and in inpatient psychiatric wards (Robertson et al., 2015). We report the commonalities across the included studies, and highlight unique findings for specific sexual minority groups, as well as any differences in findings observed across them (Popay et al., 2006).

### Fears of, and experiences of, prejudice and discrimination within mental health services

Within Talking Therapies services, primary care counselling services, and inpatient mental health wards, sexual minority service users’ feared experiencing discrimination or prejudice. In the mixed methods study by Foy et al., (2019), an unvalidated survey was used to explore the experiences of people from sexual minorities within primary care counselling and Talking Therapies services. Of 136 respondents, 57 (41.9%) had been concerned about experiencing discrimination or stigma relating to their sexual orientation, or another characteristic, in the process of accessing psychological help.

Qualitative findings highlighted that service users feared disclosing their sexual orientation due to concerns about discrimination or stereotyping by staff, and in some cases, avoided disclosing their sexual orientation as a form of self-protection (Foy et al., 2019; Morris et al., 2022). Bisexual individuals also expressed concerns that practitioners would view them as “confused, attention seeking, or hypersexual” (Morris et al., 2022). One study that conducted interviews with lesbian and gay individuals who had experience of being on an inpatient mental health ward found that all of them had anticipated experiencing homophobia from others (Robertson et al., 2015).

These fears were not always unfounded, with some service users’ reporting actual experiences of prejudice and discrimination from services (Foy et al., 2019; Morris et al., 2022; Robertson et al., 2015). In the study by Foy et al., (2019), of 136 respondents to the question, 21 (15.4%) reported experiencing prejudice or discrimination that was related to, or could have been related to, their sexual orientation at some point between trying to access support and being discharged,.

In the qualitative studies, service users from sexual minorities reported experiences of practitioners pathologizing their sexual identity. These experiences included practitioners viewing the presenting problems of service users from sexual minorities as being ‘symptomatic’ of their sexual orientation, or practitioners insisting that an individual’s sexual orientation was related to their mental health difficulties, even after the service user had stated that this was not the case (Foy et al., 2019; Morris et al., 2022). In these same settings, biphobia was the most commonly reported form of prejudice, and included stereotyping, inappropriate scrutinization of service user’s sexual identity, and being mislabelled (e.g., being called a lesbian when this was not the case; Foy et al., 2019). Within inpatient mental health wards, lesbian and gay service users’ experienced heterosexism in conversations with staff, and one participant reported experiencing homophobic sexual assault (Robertson et al., 2015).

### Inadequate clinician awareness, skills, and understanding

Some service users from sexual minorities’ perceived practitioners to lack understanding of people from sexual minorities and their unique experiences (Foy et al., 2019; Morris et al., 2022; Robertson et al., 2015). This included a lack of understanding by practitioners of: the effects of experiencing prejudice, growing up with a sexual minority identity, the higher rates of mental health concerns amongst people from sexual minorities, and the intersectionality of being both from a sexual minority and having a minoritised ethnicity (Foy et al., 2019). The potential stress of ‘coming out’ (i.e., opening up to others about one’s sexual orientation) was also noted as an area where practitioners lacked understanding (Foy et al., 2019).

Participants also reported that practitioners lacked awareness of the heterogeneity of the LGBTQ+ community, and that discrimination could also be manifested within the community itself, not just from heterosexuals (Foy et al., 2019; Morris et al., 2022). This resulted in some service users feeling misunderstood, and impacted negatively on the psychotherapeutic relationship, with participants feeling that they had to explain sexual minority related issues and experiences to practitioners (Morris et al., 2022).

### Neglecting conversations about sexual orientation

Within primary care and Talking Therapies settings, service users perceived some practitioners to be reluctant to acknowledge their sexual minority identity and its relevance to their mental health, even in instances where participants had explicitly spoken about negative events they had experienced relating to their identity (e.g., homophobia) that they felt had impacted their mental health (Foy et al., 2019; Morris et al., 2022). This omission of discussions relating to sexual orientation left participants feeling unable to explore if their sexuality contributed to how they felt about themselves (Foy et al., 2019; Morris et al., 2022). In Foy et al. (2019), when asked about whether they were asked about their sexual orientation by the service or therapist that they were referred to, 58.8% of respondents (80 of 136 participants) reported that they had not been asked.

In the context of inpatient services, lesbian and gay participants were sometimes met by silence and embarrassment by staff when they tried to talk about intimate relationships and believed that staff felt it was “taboo” for service users to discuss their sexuality. They perceived staff to be uncomfortable and lacking the skill to talk about intimate relationships or sexual orientation. This was thought to reinforce feelings of internalised homophobia (Robertson et al., 2015).

### Heteronormative assumptions and heterosexism

In primary care counselling and Talking Therapies services, people from sexual minorities had experiences with practitioners who assumed that they were heterosexual (Foy et al., 2019). These incorrect assumptions resulted in service users having to dispel them, which impeded the possibility or ease with which further discussion about sexuality could take place (Morris et al., 2022).Heteronormativity was particularly a problem for bisexual+ individuals (i.e., those with identities other than lesbian/gay; Foy et al., 2019; Morris et al., 2022) and had a profoundly negative effect on the therapeutic relationship (Morris et al., 2022). In inpatient settings, heterosexism was also experienced in conversations with staff, and in how families were referred to in the ward. This led to service users having feelings of alienation and discomfort about including reference to same-sex partners (Robertson et al., 2015).

### Positive experiences of services

People reported positive experiences when therapists were accepting, open, and non-judgmental (Foy et al., 2019). Within inpatient settings, despite an anticipation of homophobic attitudes, this was not always the reality, and in some cases, staff were supportive of same sex relationships, resulting in participants and their partners feeling welcomed and accepted (Robertson et al., 2015). One study explored individuals’ experiences of attending an LGBQ+ wellbeing group delivered by a Talking Therapies service in South London (Lloyd et al., 2021).

Attendees valued the group as providing a safe and validating space. They perceived it as normalising, helpful for allowing an understanding of the impact of minority stress, and useful for developing skills for understanding their thoughts, feelings, and behaviours. Attendees also appreciated facilitators who were open about their own sexual minority status.

### Recommendations for improving practice

52.2% of respondents (71 of 136 participants) to the survey by Foy et al., (2019) believed that services could be improved for people from sexual minorities. One recommendation for improving services was by implementing staff training, with 87.5% of respondents (119 of 136 participants) in Foy et al., (2019) believing that therapists should undergo specific training for working with people from sexual minorities (Foy et al., 2019; Morris et al., 2022).

Participants in these studies suggested that this might cover different sexualities (i.e., more than just gay/lesbian), appropriate versus inappropriate language (i.e., not assuming that words used in wider culture about the community are used in the same way in the community), the unique challenges facing sexual minorities (including coming out and experiences of discrimination), intersectionality, how gender relates (or doesn’t) to sexuality, and bisexual erasure (e.g., denying or invalidating one’s bisexual identity; Foy et al., 2019; Morris et al., 2022). In one study in these same settings, participants highlighted that whilst practitioners should be taught about commonalities across service users, this should not inadvertently reinforce generalisations about sexual minorities and their treatment needs (Foy et al., 2019). Service users believed that practitioners should be trained about the diversity of the sexual minorities, and that the training should avoid inadvertently reinforcing “sweeping generalisations” (Morris et al., 2022). They also suggested that mental health practitioners should ask where someone’s sexuality fits in to their problems, if it fits at all, rather than assuming that it plays a major role (Foy et al., 2019). Participants felt that such training should be delivered by someone who specialised in sexual minority issues, or by LGBQ+ charities (Foy et al., 2019).

Other suggestions for improving services included having health professionals with shared identities and experiences, having visible signs of inclusivity by actively showing that services are welcoming of people from sexual minorities and equipped to help them, in the form of staff and services (including on-line services) that specialise in LGBQ+ issues, (Morris et al., 2022).

### Treatment outcomes and negative effects of psychological treatment

Four of the included quantitative studies explored the treatment outcomes of adults from sexual minorities in statutory mental health services, all of which were in the context of Talking Therapies services. One of these studies specifically explored outcomes of adults from sexual minorities who had attended a wellbeing group specifically for people from sexual minorities at a Talking Therapies service in South London (Hambrook et al., 2022). The intervention was based on CBT and had been developed specifically for people from sexual minorities, and there was evidence that symptoms of depression, anxiety, and functional impairment improved between the beginning and end of the intervention.

Three of the other quantitative studies explored treatment outcomes in relation to routine care received in Talking Therapies services (Kent et al., 2025; Rimes et al., 2018, 2019). The study by Rimes et al., (2018) used data from four services provided by a large South London mental health Trust and found that both lesbian and bisexual women had smaller reductions in depression and functional impairment symptoms than heterosexual women. Bisexual women also had smaller reductions in anxiety symptoms and were more likely to not meet the criteria for recovery from depression and anxiety, and more likely to not meet the criteria for reliable recovery for depression and anxiety. No differences were found between bisexual and gay men relative to heterosexual men.

These results are supported the study by Rimes et al., (2019) which used data from all Talking Therapies services in England found that, relative to heterosexual women, bisexual and lesbian women had higher final session scores for anxiety, depression, and functional impairment, and were at increased risk of not meeting reliable recovery for anxiety/depression and functional impairment. Consistent with the study by Rimes et al., (2018), no differences were observed between gay men and heterosexual men, but in contrast, bisexual men also had had higher final session scores for depression and functional impairment, and were at increased risk of not meeting reliable recovery for anxiety/depression and functional impairment relative to heterosexual and gay men.

Kent et al., (2025) used routine data from seven Talking Therapies services in North and Central East London, and had a considerably larger sample of sexual minorities (n = 7422 relative to n = 1130 in Rimes et al., (2018) and n = 4472 in Rimes et al., (2019)). Kent et al., (2025) found that gay men had greater odds of meeting the criteria for reliable improvement and reliable recovery for depression/anxiety and also showed significantly greater improvements in depression and anxiety severity, relative to heterosexual men. No differences were found between any other sexual minority groups and heterosexuals. The results also showed that gay and bisexual men, and bisexual women, had lower odds of attrition from psychological therapy relative to heterosexual men and women. No differences were observed between lesbian and heterosexual women.

One of the included studies explored the perceived negative effects from psychological therapy for common mental disorders in primary and secondary care, and found that individuals who identified as bisexual had greater odds of self-reporting that they had experienced long-lasting negative effects from psychological therapy.

### Young People from Sexual Minorities experiences

Two of the included studies explored young people from sexual minorities experiences of receiving Dialectical Behavioural Therapy (DBT) in a national and specialist DBT service provided by NHS Child and Adolescent Mental Health Services (CAMHS). One of these explored clinical outcomes across sexual minority groups (including heterosexuals) and found that all groups benefited from the intervention (Camp et al., 2024).

The other study was qualitative and explored the experiences of young people from sexual minorities accessing the service (Camp et al., 2023). Young people generally found receiving DBT in the service helpful, with them highlighting that some of the skills learned in DBT could be helpful for coping with minority stressors (e.g., using mindfulness of thoughts to diffuse from homophobic thoughts). Participants reported feeling accepted, supported, and non-judged, and generally felt that therapists were open and less prone to making assumptions than previous service experiences, which helped the space to feel safe. However, some young people reported that DBT sometimes did not allow enough space for exploring sexual identity issues, and there was a lack of attention paid applying DBT skills to sexual identity issues which could act as a barrier to DBT addressing their minority-related needs.

## Discussion

This review synthesised the available evidence on the experiences of sexual minority users of statutory mental health services in the UK. We found only 11 studies that met our inclusion criteria, predominantly with adults, and with the majority in primary care counselling and Talking Therapies services, suggesting that this is an under researched area.

Following guidance by Popay et al. (2006), we begin with a discussion of the commonalities and differences that we observed across the included studies. Despite the results of the review relying on a relatively small number of studies, they were generally of good quality, and findings were mostly consistent from the studies which were set in Talking Therapies services, Primary Care counselling services, and inpatient settings. They show that some service users from sexual minorities are reporting negative experiences in services that are related to their sexual identity, despite protections being in place for sexual minorities since 2010 that mean that experiences, such as discrimination, should not be occurring within services at all (The Equality Act, 2010).

Across these settings, people from sexual minorities described the anticipation of experiencing discrimination or prejudice from staff, even in the absence of any indicators that practitioners might behave in this way. Bisexual individuals in particular had concerns that practitioners would express stereotypical views, such as thinking that the individual was “confused” or “hypersexual” (Morris et al., 2022). This led to some concealing their sexual orientation or selective disclosure. Foy et al., (2019), found that a large proportion (41.9%) of respondents reported this kind of anticipatory anxiety in the process of accessing psychological support in primary care.

The included studies also demonstrated that these fears were appropriate. The qualitative evidence, and evidence from the quantitative component of the mixed methods study, corroborated that many individuals had negative experiences in mental health services ranging from so called microaggressions (i.e., intentional or unintentional brief and commonplace slights or invalidations based on someone’s identity; (Nadal, (2013)) in the form of heteronormative assumptions, to the explicit pathologisation of their sexual orientation. In one study biphobia was the most commonly reported type of prejudice reported (Foy et al., 2019).

Although it is unclear from the included studies whether such behaviour by clinicians is as a result of personal attitudes and/or beliefs, or other factors, the qualitative research we identified highlighted that a lack of knowledge and skills among practitioners may have a role to play. This is despite recommendations from the NHS England Equality and Diversity Council that there should be mandatory training for staff on working with LGBT+ patients (NHS, 2015). In both primary care and inpatient settings, some service users perceived practitioners to lack an understanding of different sexual minority identities and the diversity of the LGBQ+ population. There were also concerns that staff had little or no awareness of the likely stressors for people from sexual minorities, many of which may contribute to mental ill-health such as what it is like to grow up LGBQ+, coming out, and to experience prejudice and discrimination.

These experiences were reported to undermine the therapeutic relationship, and in some instances, deprived individuals of the opportunity to explore whether and how their sexual orientation may relate to their mental health difficulties.

The findings of this review are in line with systematic reviews of the international literature on this topic that have also found that sexual minorities report experiences with practitioners who pathologize their identity, make heteronormative assumptions, and lack knowledge about working with LGBQ+ people (McNamara & Wilson, 2020; River et al., 2025).

We found notably conflicting findings across the three quantitative studies that investigated treatment outcomes in routine care from Talking Therapies services (Kent et al., 2025; Rimes et al., 2018, 2019). The study by Kent et al., (2025) found that gay men had more favourable treatment outcomes relative to heterosexual men, but found no differences between other sexual minority groups relative to heterosexuals. Whilst the results of this study are convincing, due to the studies high quality and the largest sample of sexual minority participants relative to the other two studies, it is noteworthy that the study only used data from services in North and Central East London. As acknowledged in the study, North and East London has the highest proportion of sexual minorities in England, and therefore increased opportunities for contact and support from other sexual minorities may act as a protective factor for minority stress levels which may partly explain the observed findings. Additionally, the significant differences across outcomes for gay men relative to heterosexual men were small (e.g., on average gay men had a reduction of 0.51 points greater than that of heterosexual men for depression severity) and were unlikely to reflect a clinically important difference (Kent et al., 2025).

In contrast to the study by Kent et al., (2025), the study using data from all Talking Therapies services across England found that bisexual individuals and lesbian woman had worse outcomes relative to heterosexuals, but no differences were observed between gay and heterosexual men (Rimes et al., 2019). Further research is needed, with a large sexual minority sample, to understand whether a treatment disparity exists between certain sexual minority groups and heterosexuals, and whether this varies by geographical location.

None of the included quantitative studies investigated potential mechanisms that might influence treatment outcomes for certain sexual minority groups. However, the findings of this review may help to identify some of the factors that might contribute to these potentially poorer outcomes. It is possible that the minority stressors experienced in everyday life are reinforced within primary care and inpatient services for some sexual minorities. There is evidence that experiencing minority stressors in general (i.e., not specifically in services) is associated with worse mental health outcomes for sexual minorities (Hatzenbuehler et al., 2009; Lee et al., 2016; Marchi et al., 2024). It is therefore possible that experiencing such stressors within services (e.g., anticipating discrimination, pathologization of sexual identities) may contribute to poorer outcomes, although we cannot conclude this from our included studies. However, relative to gay/lesbian individuals, bisexuals are more likely to experience minority stress in the form of stigma, victimization, invisibility/erasure, and identity concealment (Dodge et al., 2016; Ross et al., 2018). Women from sexual minorities are also more likely to report childhood trauma and interpersonal violence in adulthood, in addition to gender-based prejudice and discrimination throughout the life course, than heterosexual women (Austin et al., 2008; Platt et al., 2016; Szalacha et al., 2017). Evidence also suggests that daily experiences of heterosexism contribute to the maintenance of PTSD symptoms in women from sexual minorities (Dworkin et al., 2018). A combination of these and other factors may therefore contribute to the differences in outcomes we identified in two of the included studies for bisexual individuals and lesbian women (Rimes et al., 2018, 2019), particularly if services are failing to address the minority stressors that are contributing to and/or maintaining mental health difficulties.

Future research should adopt longitudinal study designs and systematically measure whether negative experiences in services mediate the relationship between sexual orientation and treatment outcomes and establish the pathways by which this occurs.

For example, the therapeutic alliance is associated with more favourable psychotherapeutic outcomes, with the alliance possibly being a mediator of change underlying service users’ response to psychotherapy (Baier et al., 2020; Cameron et al., 2018). Future research might explore whether the negative experiences reported by sexual minority users of mental health services results in a poorer therapeutic alliance which may consequently result in poorer outcomes.

Importantly, some positive experiences of mental health care were reported in both primary care and inpatient settings, where practitioners were open, non-judgmental, accepting, and affirming of same-sex relationships.

It is noteworthy that the included qualitative studies may have been more likely to attract individuals who have had a negative experience with services leading to response bias. As a result, the included studies are predominantly weighted towards negative experiences. Nevertheless, the value of having a safe and affirming space to explore the impact of minority stress on the mental health of attendees at a wellbeing group specifically for sexual minorities was clear from two of our included studies (Hambrook et al., 2022; Lloyd et al., 2021). This suggests that having LGBQ+ specific groups might be valuable for sexual minorities, or having professionals deliver therapeutic interventions that are adapted to consider the role of minority stressors. A model exists for adapting evidence-based treatments to be LGBQ+ affirmative by adopting minority stress principles (Pachankis et al., 2023), and there is some evidence that adapting intervention in this way may be a promising avenue.

A systematic review of CBT, and its adaptations, aimed at improving the mental health of sexual (and gender) minorities found that the included studies generally showed positive findings (Tudor-Sfetea & Topciu, 2024). However, the review also highlighted that there were significant methodological limitations of the included studies (e.g., due to issues with confounding and the measurement of outcomes), meaning that further research is needed.

We only found two studies that explored the experiences of sexual minority young people, and both were about young people’s experiences of a national and specialist DBT CAMHS service. This highlights a significant gap in the literature which is concerning in light of the increased psychological burden experienced by this group. Finally, turning to the experiences of sexual minority staff, we found no research that had explored their experiences within statutory mental health services in the UK, meaning that we couldn’t answer our second research question. It is therefore unclear what the experiences of working in statutory mental health services are for staff from sexual minorities in the UK. However, as highlighted in the introduction, the Royal College of Psychiatrists conducted a survey of its members in 2021/22 and explored the experiences of sexual and gender minority (i.e., LGBTQ+) psychiatrists (Royal College of Psychiatrists, 2022). The survey found that 48% had experienced bullying, harassment, and microaggressions within the preceding three years, with microaggressions being the most common experience.

These findings suggest that mental health professionals could be having negative experience in mental health services that are related to their sexuality. However, as this survey included sexual and gender minorities, as well as psychiatrists working in non-NHS settings and overseas, firm conclusions can’t be reached about the experiences of sexual minority mental health professionals more broadly, or of their experiences in statutory mental health services in the UK.

## Implications

The findings of this review demonstrate that the attitudes and biases held by society can and do find their way into mental health services (King, 2015), and that more needs to be done to improve the experiences of people from sexual minorities accessing statutory mental health services in the UK.

One potential avenue suggested by our findings for improving the experiences for sexual minorities is by providing training for NHS mental health staff to ensure a consistent level of cultural competence in working with this group. There is recognition that current standard Equality, Diversity, and Inclusion training that all NHS staff receive is not enough to articulate the needs and experiences of sexual minorities (Truscott et al., 2022). Additionally, within Talking Therapies services, there is evidence that the amount of sexual orientation training that clinicians receive, both pre- and post-qualification, significantly varies in length and depth, with some clinicians reporting receiving no training at all (Ho et al., 2023).

There have been recommendations and commitments made from both the UK Government and the NHS to increase the awareness of health professionals about LGBTQ+ experiences in order to improve patient care, and a commitment by the government to embed LGBTQ+ issues in mental health services (Government Equalities Office, 2018; Truscott et al., 2022; NHS England, n.d.,). The NHS Confederation has also developed a practical framework, the Health and Care LGBTQ+ Inclusion Framework, aimed at health and social care leaders to enable them to create more inclusive environments for LGBTQ+ service users and staff (Truscott et al., 2022). One of the pillars of inclusivity in the framework is that staff have a strong knowledge base, which includes staff understanding the specific needs of sexual (and gender) minorities and the inequalities that they face.

Preliminary research suggests that training might be a promising avenue for improving health professionals knowledge, skills, and attitudes when working with sexual and gender minorities in healthcare more broadly (Damery et al., 2025; Sekoni et al., 2017; Yu et al., 2023). Two of the studies included in this review also highlight the potential value of training mental health professionals to deliver therapeutic groups, specifically for sexual minorities, that incorporate principles of Minority Stress Theory (Hambrook et al., 2022; Lloyd et al., 2021).

However, training in and of itself is not enough. As highlighted by the included studies, some sexual minorities fear experiencing prejudice and discrimination within services, even in the absence of any indicators that this might occur. They highlighted that services and clinicians need to show visible signs of inclusivity to help ease any concerns about discrimination, and to demonstrate that services are welcoming of sexual minorities and that clinicians are adequately trained to work with sexual minorities and their experiences (Morris et al., 2022).

The included studies also highlight the importance of routinely collecting sexual orientation data in health services (Kent et al., 2025; Rimes et al., 2018, 2019). The collection of sexual orientation in Talking Therapies services has allowed the elucidation of potentially poorer treatment outcomes for lesbian women and bisexuals. Despite the Sexual Orientation Monitoring Information Standard, a mechanism for healthcare services in England to record the sexual orientation of service users aged 16 and over, being introduced by NHS England in 2017 (NHS England, 2017), there remain challenges for healthcare providers in consistently collecting this information, including inadequate data collection systems, a lack of awareness by staff of the health inequalities experienced by sexual minorities, and staff feeling uncomfortable about asking service users. about their sexual orientation (Truscott., 2022; Pollard et al., 2019). Sexual orientation monitoring needs to be scaled up and consistently applied across the NHS, in all parts of the UK, and this data should be used to build the evidence base for the experiences and outcomes of sexual minorities in (mental) health services.

To further understand how services can best support people from sexual minorities, more research is needed to determine whether treatment inequalities exist between certain sexual minority groups and heterosexuals in Talking Therapies services, and the potential mechanisms that influence any disparities, as well as exploring whether inequalities exist in other mental health service settings. Whilst the included qualitative studies highlight the stressors that sexual minorities are experiencing, longitudinal studies are needed to understand the pathways by which experiences of minority stressors (both out with and in services) are related to treatment outcomes. However, it is essential that positive change is not delayed until we reach that understanding. The studies in this review highlight that sexual minorities are reporting experiences of prejudice and discrimination in services, and these experiences should not be occurring at all.

Finally, research is needed to understand the experiences of sexual minority staff, sexual minority young people, and the experiences of sexual minorities in different statutory mental health settings (i.e., beyond primary care counselling and Talking Therapies services).

## Strengths and Limitations

We first report on the robustness of the synthesis by reflecting on the method of synthesis used and its limitations, the quality, validity, and generalisability of the included studies, and any discrepancies or uncertainties identified (Popay et al., 2006). We followed guidance as by Popay et al., (2006) and conducted a preliminary synthesis by tabulating the findings of the included studies. This was used to identify commonalities across studies, service contexts, and populations, and to identify any unique findings for specific sexual minority groups. We also highlighted any notable differences in findings and critically reflected on whether this was related to the study design, or other variables.

The included studies were generally of good quality meaning they are generally reliable sources of evidence. Results were also largely consistent across studies and service contexts, increasing the reliability of the results and conclusions of this review. However, there are some issues with generalisability, and some uncertainties were identified. Whilst the included qualitative studies, and the mixed methods study, consistently found that people from sexual minorities are having negative experiences within services that is related to their sexual identity, the use of an unvalidated questionnaire, and issues with the sampling strategy and relatively small sample size in the mixed methods study mean that the percentages reported throughout the results should be considered with caution, and should not be seen as reflective of the true proportion of sexual minorities having these experiences within services.

In relation to uncertainties identified in the review, the key uncertainty is whether some sexual minority groups have poorer treatment outcomes. It is possible that there may be geographical variation in treatment inequalities, but research is needed to explore this possibility.

In relation to the strengths and limitations of the conduct of this review, we adopted a robust search strategy by consulting with a subject specialist librarian at our institution in the development of the search terms and strategy which ensured that we had the best chance of identifying all eligible literature.

We also ensured a stringent data extraction and quality assessment process, with a second reviewer independently conducting full text screening, extracting data and conducting quality assessments of included studies.

There are some limitations to acknowledge. Our research question was quite broad (i.e., what are the “experiences” of services). This is partly due to the lack of universally accepted formal conceptualisation of what “patient experience” means (see Oben, 2020) and it was therefore challenging to develop specific key terms to capture this concept which means that some relevant publications could have been missed. However, we developed our key search terms based on studies we were aware of that we believed would meet our eligibility criteria and piloted the search strategy to ensure that these papers were captured in our search. We also made use of MeSH terms/Subject Headings to identify relevant literature.

Finally, this review only explored the experiences of people from sexual minorities and precludes any conclusions or inferences about the experiences of those with intersecting minority identities (e.g., gender diverse individuals who also identify as a sexual minority). Whilst people from gender minorities also report negative experiences in mental health services related to their gender identity (Ellis et al., 2015), conflating sexual orientation and gender identity has been criticised as it can risk overlooking the unique experiences of both groups (Ho et al., 2023).

## Conclusion

Some sexual minority service users have negative experiences within statutory mental health services in the UK that are related to their sexual identity. These include anticipatory anxiety about experiencing prejudice and discrimination in services, microaggressions (e.g., heteronormative assumptions) and pathologization of sexual minority identities. There are mixed findings about whether certain sexual minority groups have worse outcomes in Talking Therapies services. In line with UK government, NHS, and sexual minority service users’ recommendations, mental health services should consider introducing training for all staff to improve their competence in working with people from sexual minorities.

## Data Availability

N/A

## Appendix I Key Words used in the Bibliographic Searches

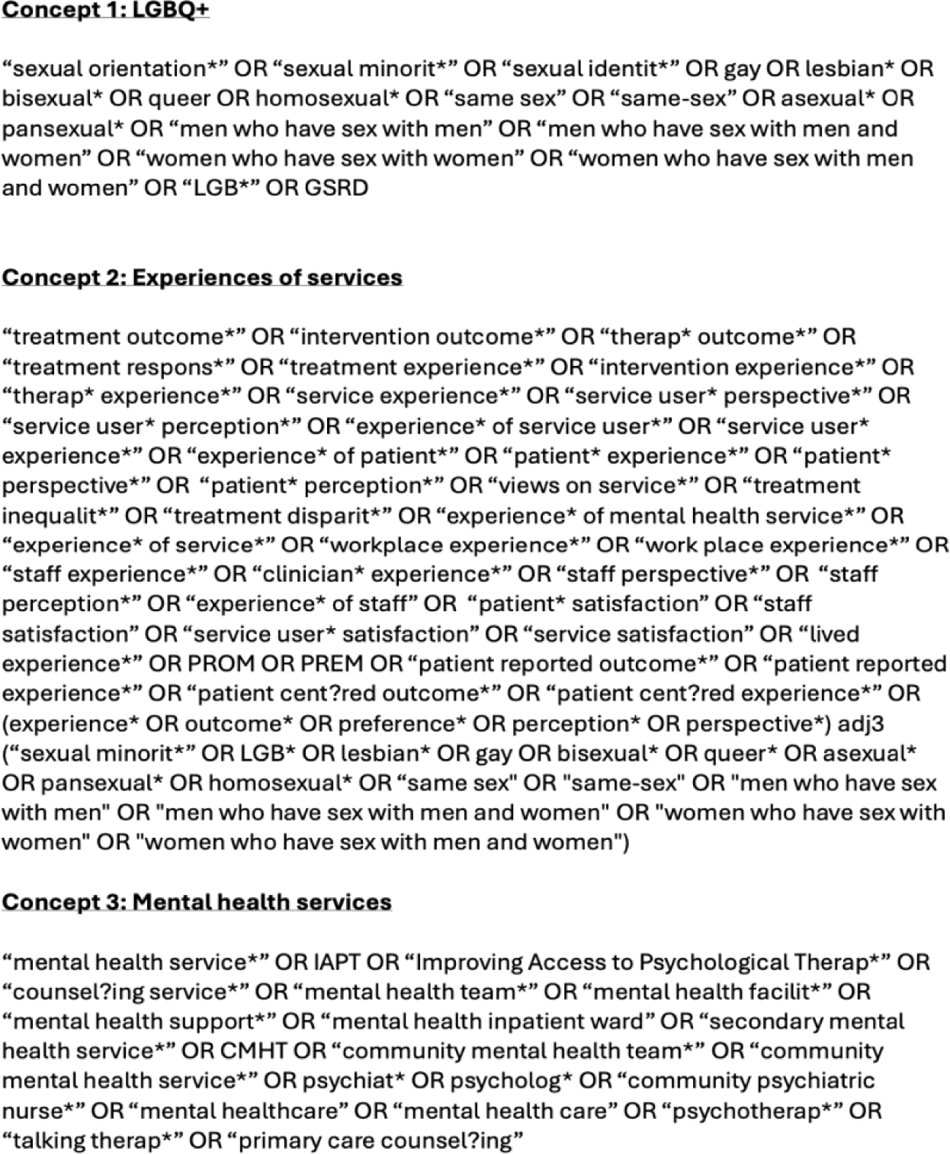

## Appendix II Quality Assessment for the Included Quantitative Studies

Quality Assessment for the Included Quantitative Studies
|  | Comparison of Treatment Outcomes Between Lesbian, Gay, Bisexual and Heterosexual Individuals Receiving a Primary Care Psychological Intervention (Rimes et al., 2018) | Sexual Orientation Differences in Psychological Treatment Outcomes for Depression and Anxiety: National Cohort Study (Rimes et al., 2019) | Group intervention for sexual minority adults with common mental health problems: preliminary evaluation (Hambrook et al., 2022) | Patient experience of negative effects of psychological treatment: results of a national survey (Crawford et al., 2016) | Clinical outcomes for sexual and gender minority adolescents in a dialectical behaviour therapy programme (Camp et al., 2024) | Psychological therapy outcomes by sexual orientation and gender: a retrospective cohort study (Kent et al., 2025) | LGBQ+ adults' experiences of Improving Access to Psychological Therapies and primary care counselling services: informing clinical practice and service delivery (Foy et al., 2019) |
| --- | --- | --- | --- | --- | --- | --- | --- |
| 1. Question/objective sufficiently described? | 2 | 2 | 2 | 2 | 2 | 2 | 2 |
| 2. Study design evident and appropriate? | 2 | 2 | 2 | 2 | 2 | 2 | 2 |
| 3. Method of subject/comparison group selection or source of information/input variables described and appropriate? | 2 | 2 | 2 | 1 | 2 | 2 | 1 |
| 4. Subject (and comparison group, if applicable) | 1 | 1 | 1 | 2 | 2 | 2 | 2 |
| characteristics sufficiently described? |  |  |  |  |  |  |  |
| 5. If interventional and random allocation was possible, was it described? | N/A | N/A | N/A | N/A | N/A | N/A | N/A |
| 6. If interventional and blinding of investigators was possible, was it reported? | N/A | N/A | N/A | N/A | N/A | N/A | N/A |
| 7. If interventional and blinding of subjects was possible, was it reported? | N/A | N/A | N/A | N/A | N/A | N/A | N/A |
| 8. Outcome and (if applicable) exposure measure(s) well defined and robust to measurement / misclassification bias? Means of assessment reported? | 2 | 2 | 2 | 0 | 2 | 2 | 1 |
| 9. Sample size appropriate? | 2 | 2 | 2 | 2 | 1 | 2 | 1 |
| 10. Analytic methods described/justified and appropriate? | 2 | 2 | 2 | 2 | 2 | 2 | 2 |
| 11. Some estimate of variance is reported for the main results? | 2 | 2 | 1 | 2 | 2 | 2 | N/A |
| 12. Controlled for confounding? | 2 | 2 | 0 | 1 | 2 | 2 | N/A |
| 13. Results reported in sufficient detail? | 2 | 2 | 2 | 2 | 2 | 2 | 2 |
| 14. Conclusions supported by the results? | 2 | 2 | 2 | 1 | 2 | 2 | 2 |
| Summary Score | 95.5 | 95.5 | 81.8 | 77.3 | 95.5 | 100 | 83.3 |

## Appendix III Quality Assessment for the Included Qualitative Studies

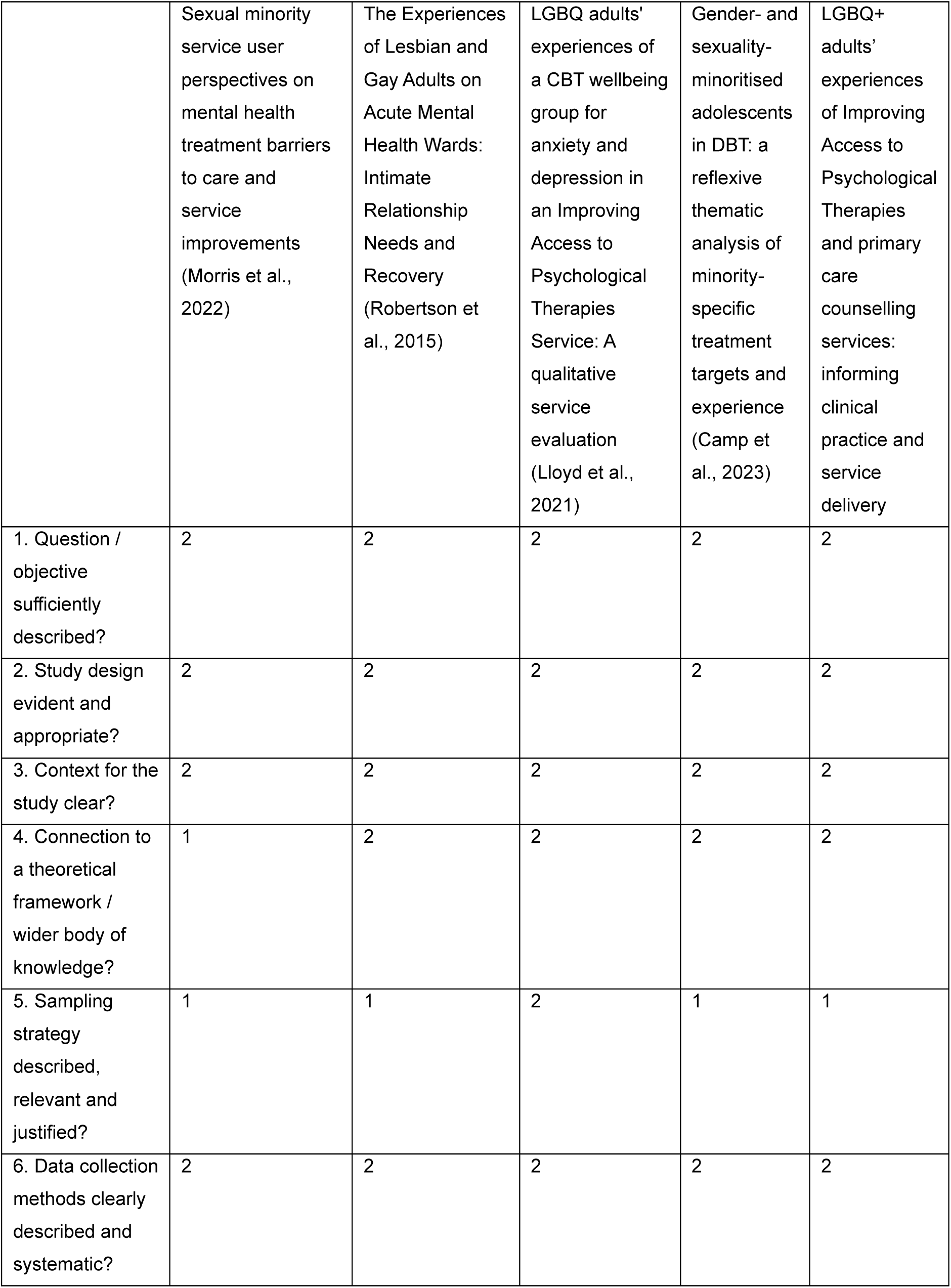
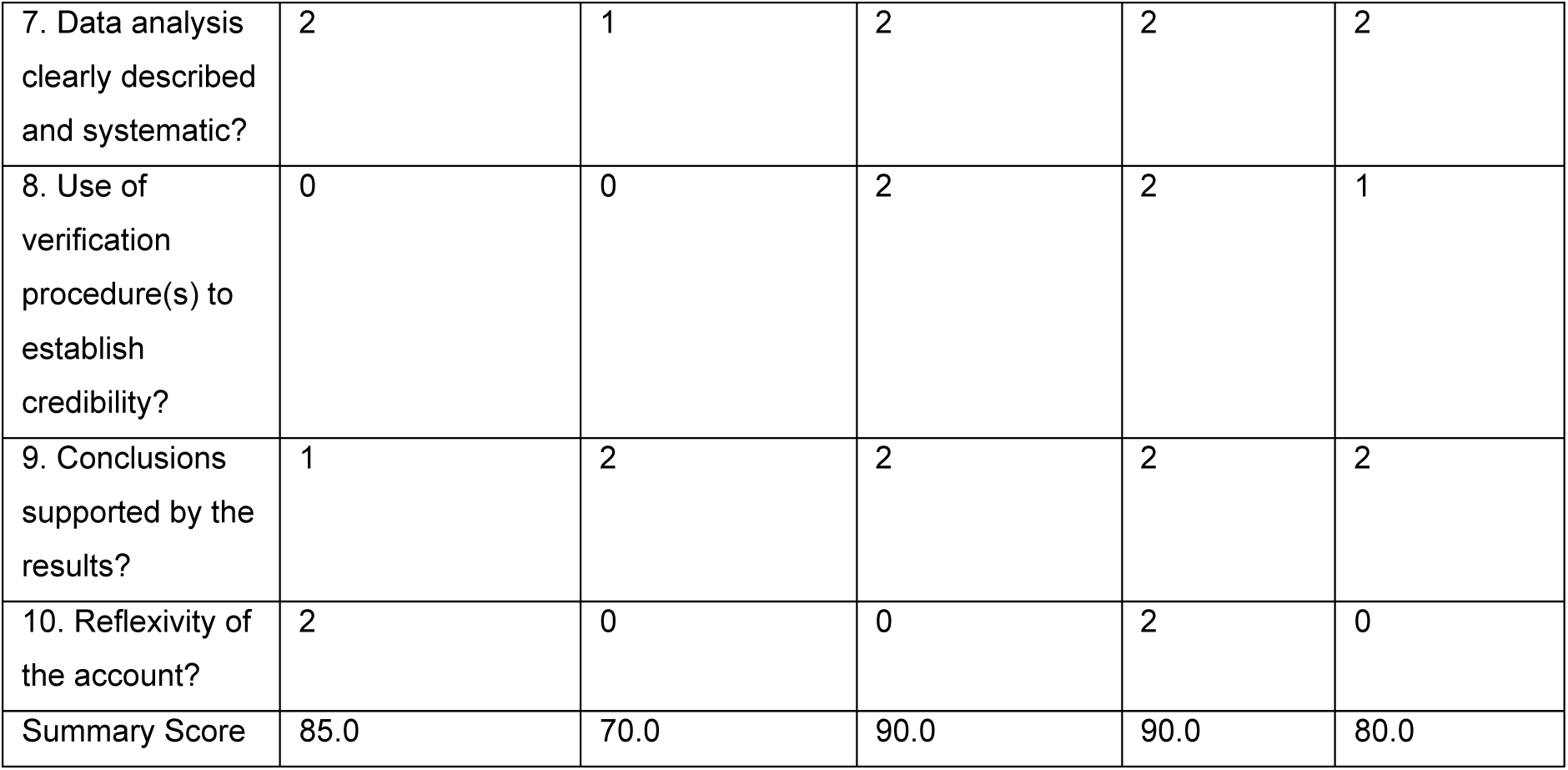

|  | Sexual minority service user perspectives on mental health treatment barriers to care and service improvements (Morris et al., 2022) | The Experiences of Lesbian and Gay Adults on Acute Mental Health Wards: Intimate Relationship Needs and Recovery (Robertson et al., 2015) | LGBQ adults' experiences of a CBT wellbeing group for anxiety and depression in an Improving Access to Psychological Therapies Service: A qualitative service evaluation (Lloyd et al., 2021) | Gender- and sexuality-minoritised adolescents in DBT: a reflexive thematic analysis of minority-specific treatment targets and experience (Camp et al., 2023) | LGBQ+ adults' experiences of Improving Access to Psychological Therapies and primary care counselling services: informing clinical practice and service delivery |
| --- | --- | --- | --- | --- | --- |
| 1. Question / objective sufficiently described? | 2 | 2 | 2 | 2 | 2 |
| 2. Study design evident and appropriate? | 2 | 2 | 2 | 2 | 2 |
| 3. Context for the study clear? | 2 | 2 | 2 | 2 | 2 |
| 4. Connection to a theoretical framework / wider body of knowledge? | 1 | 2 | 2 | 2 | 2 |
| 5. Sampling strategy described, relevant and justified? | 1 | 1 | 2 | 1 | 1 |
| 6. Data collection methods clearly described and systematic? | 2 | 2 | 2 | 2 | 2 |
| 7. Data analysis clearly described and systematic? | 2 | 1 | 2 | 2 | 2 |
| 8. Use of verification procedure(s) to establish credibility? | 0 | 0 | 2 | 2 | 1 |
| 9. Conclusions supported by the results? | 1 | 2 | 2 | 2 | 2 |
| 10. Reflexivity of the account? | 2 | 0 | 0 | 2 | 0 |
| Summary Score | 85.0 | 70.0 | 90.0 | 90.0 | 80.0 |

## Appendix IV Summary of Findings for the Included Studies

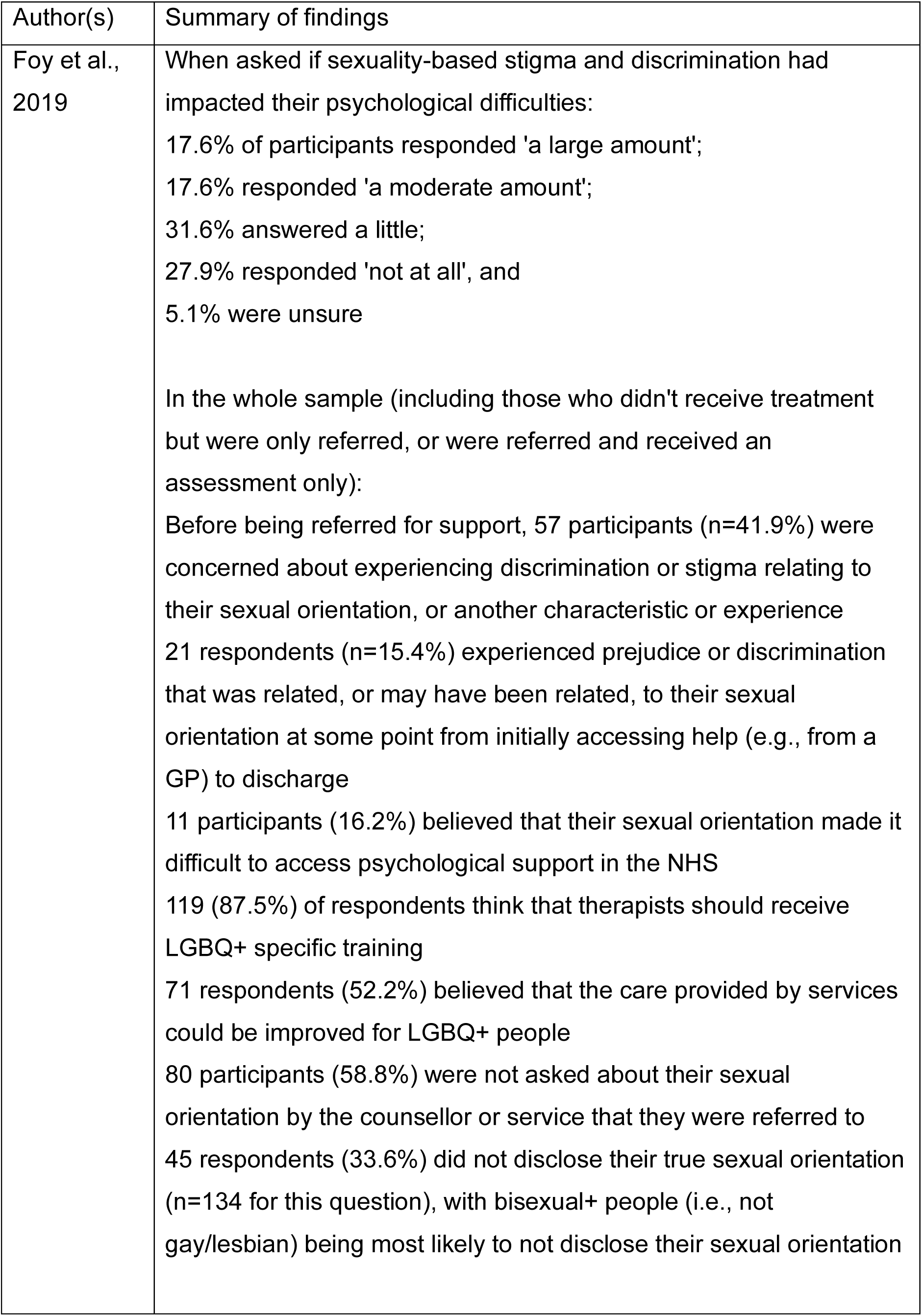
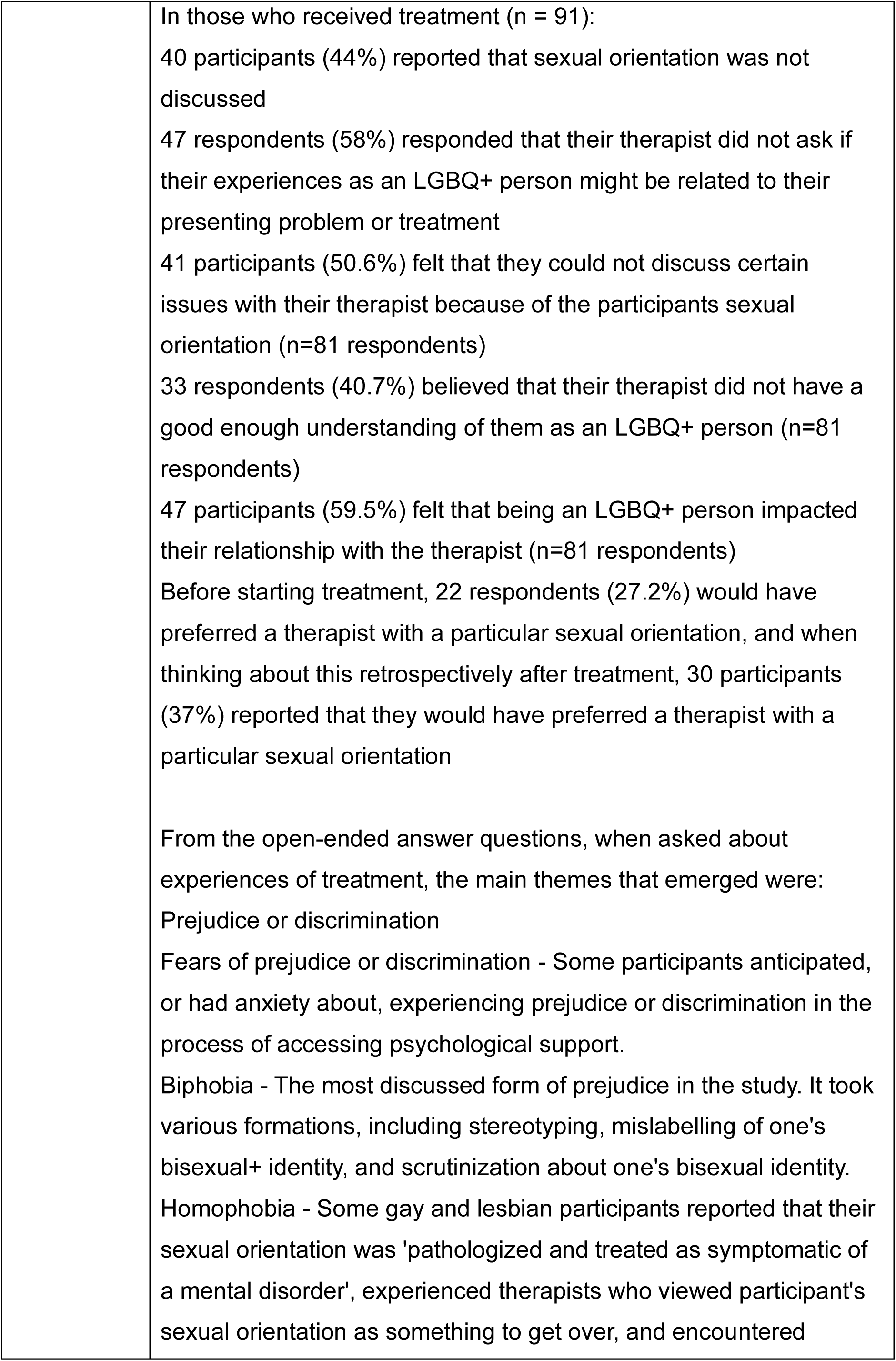
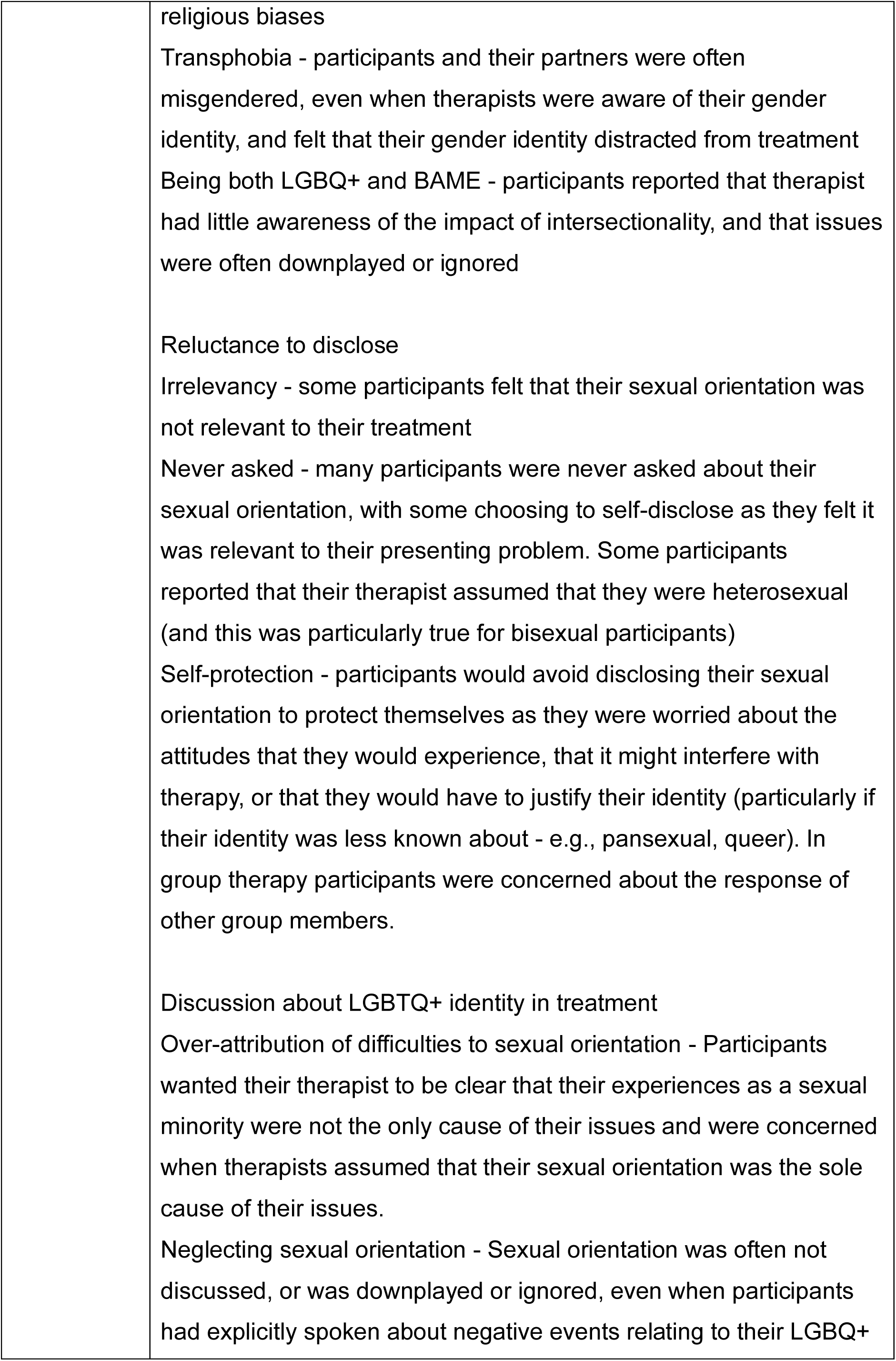
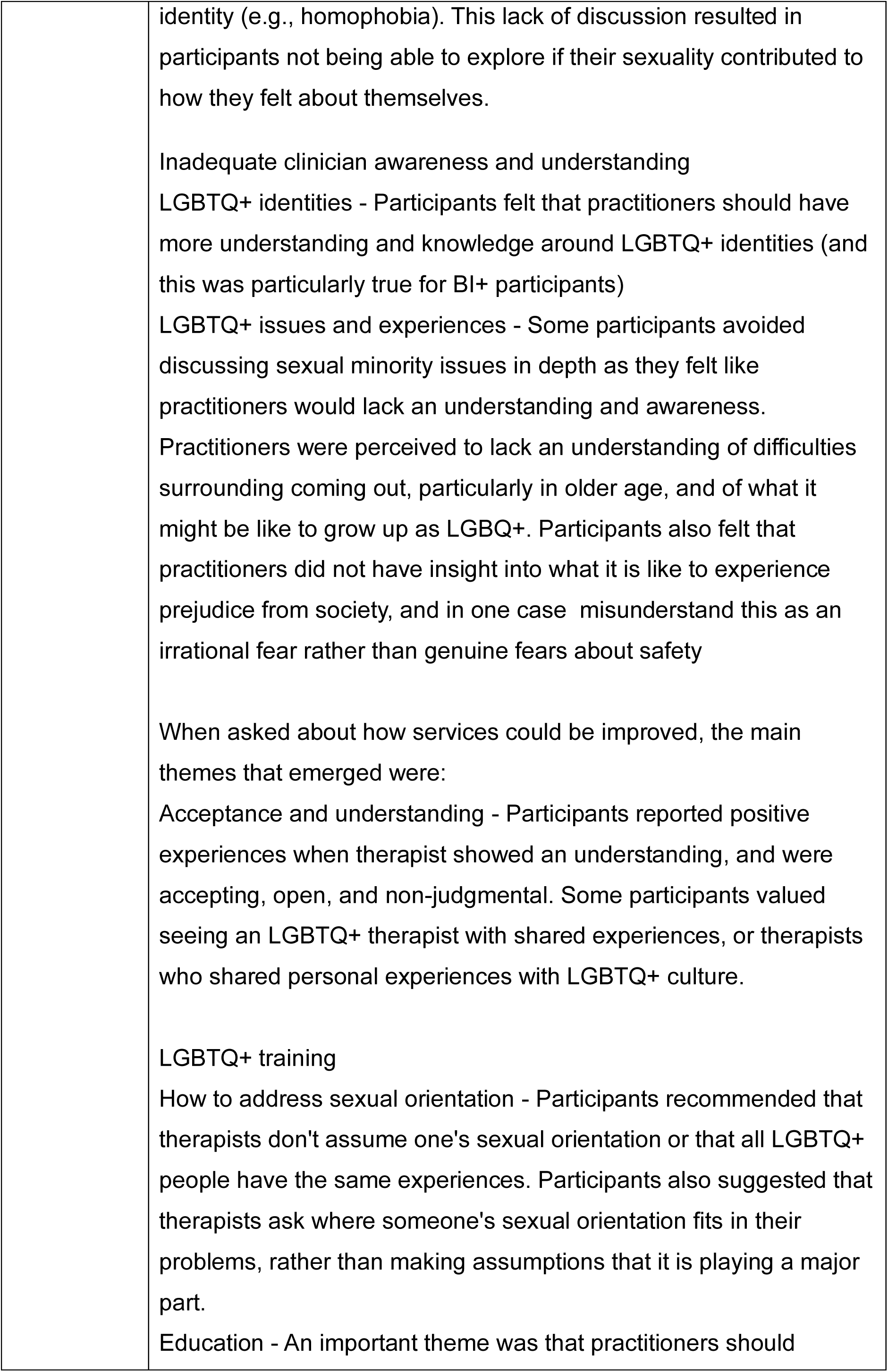
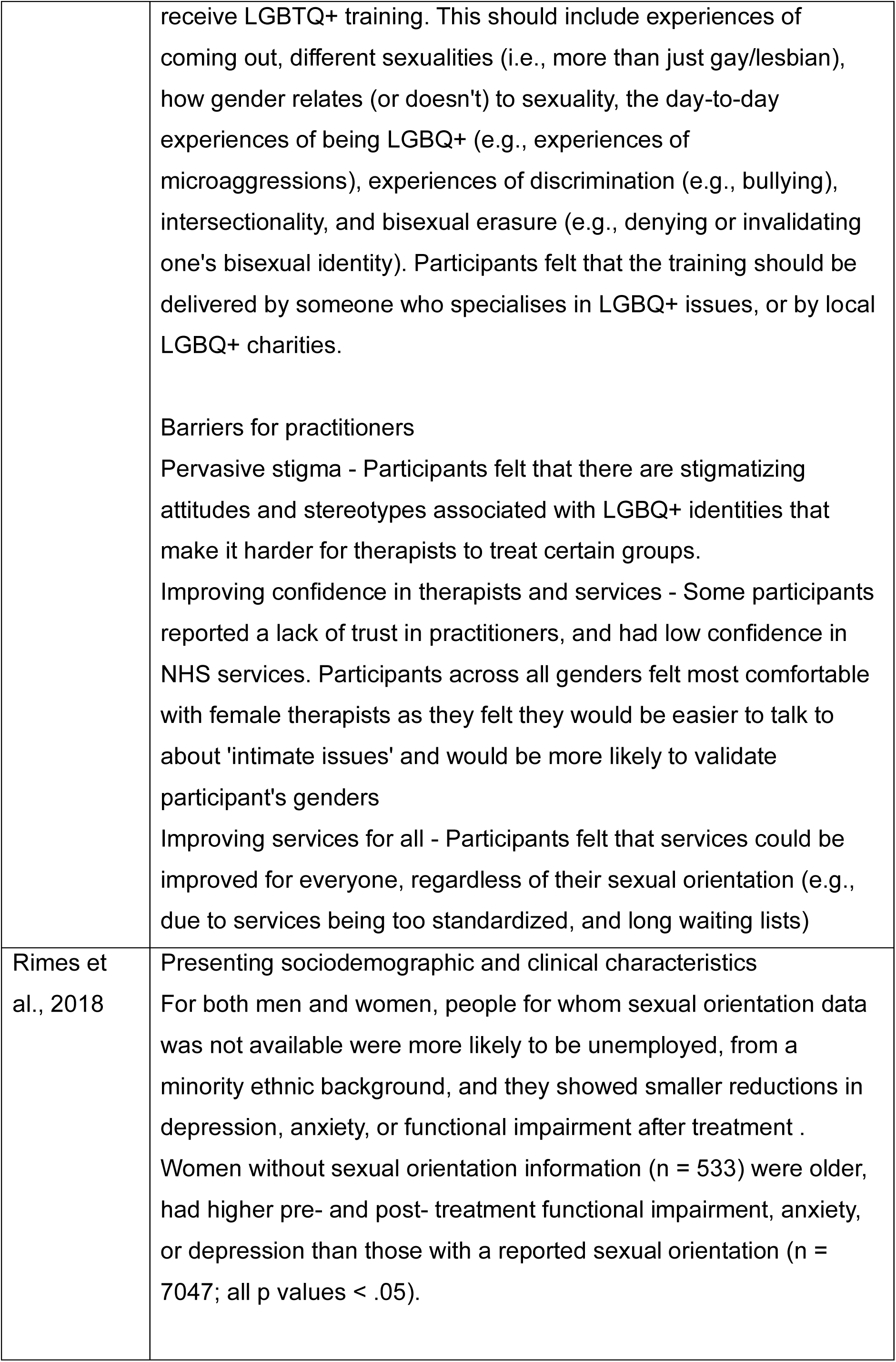
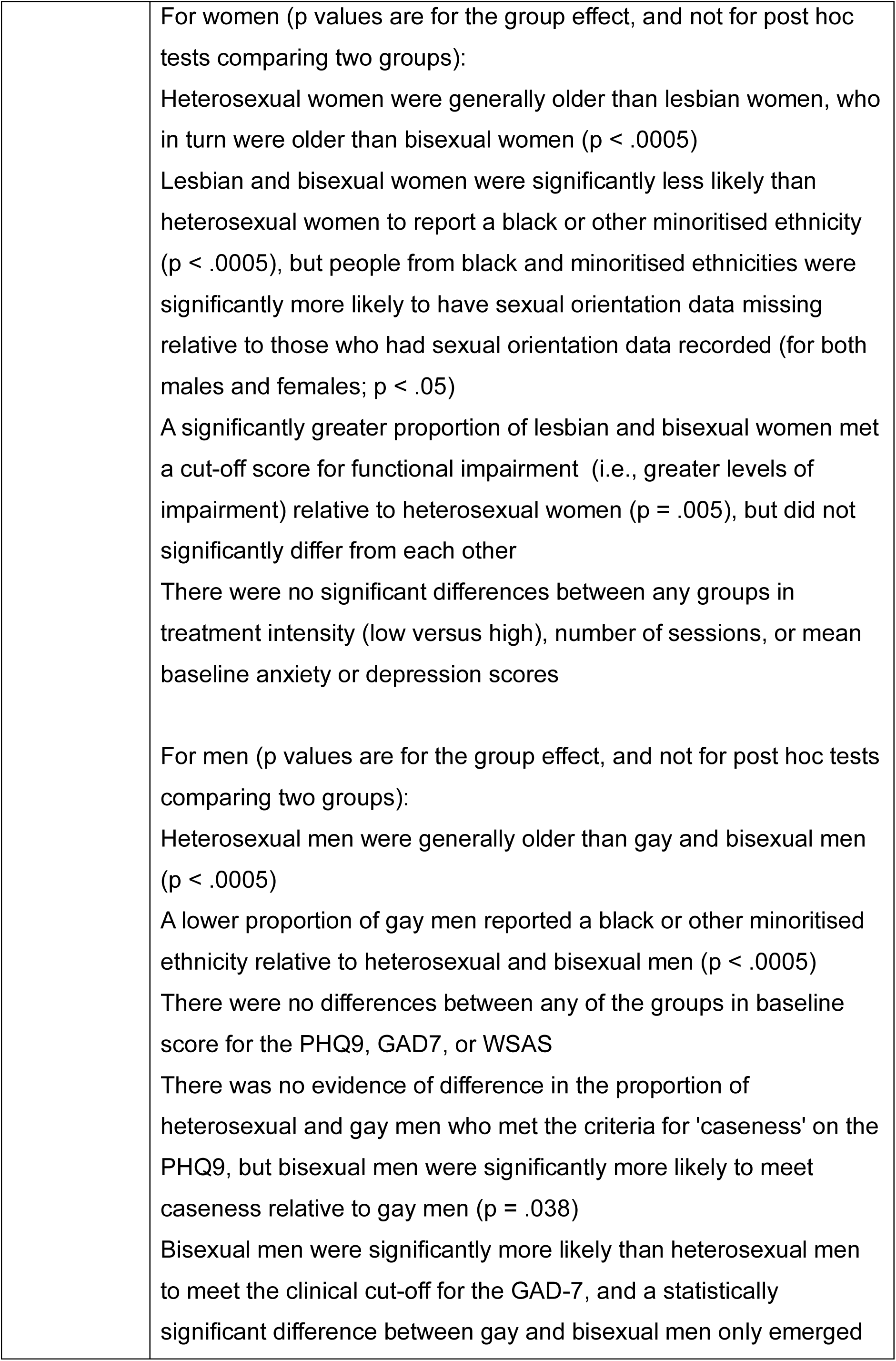
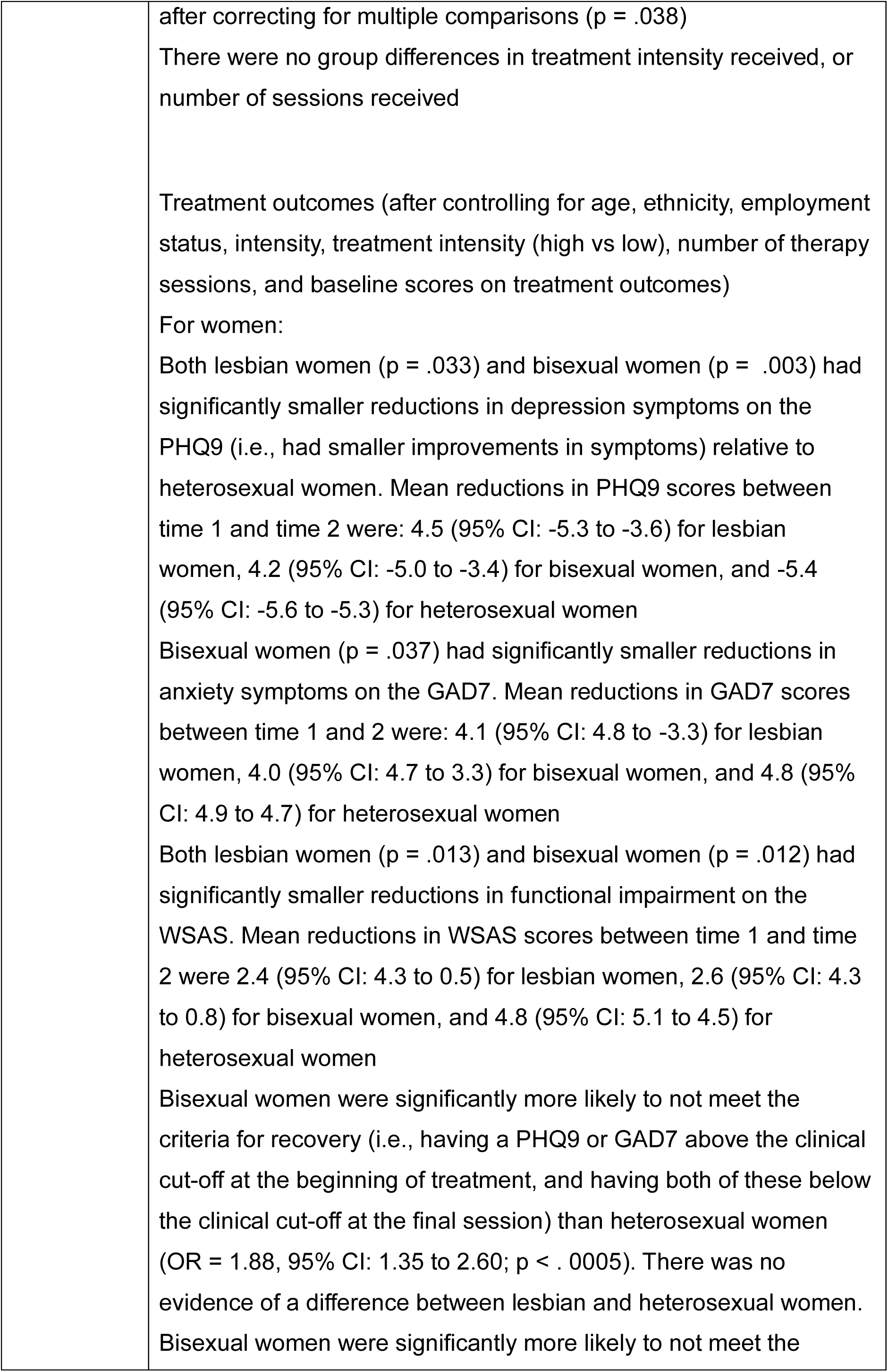
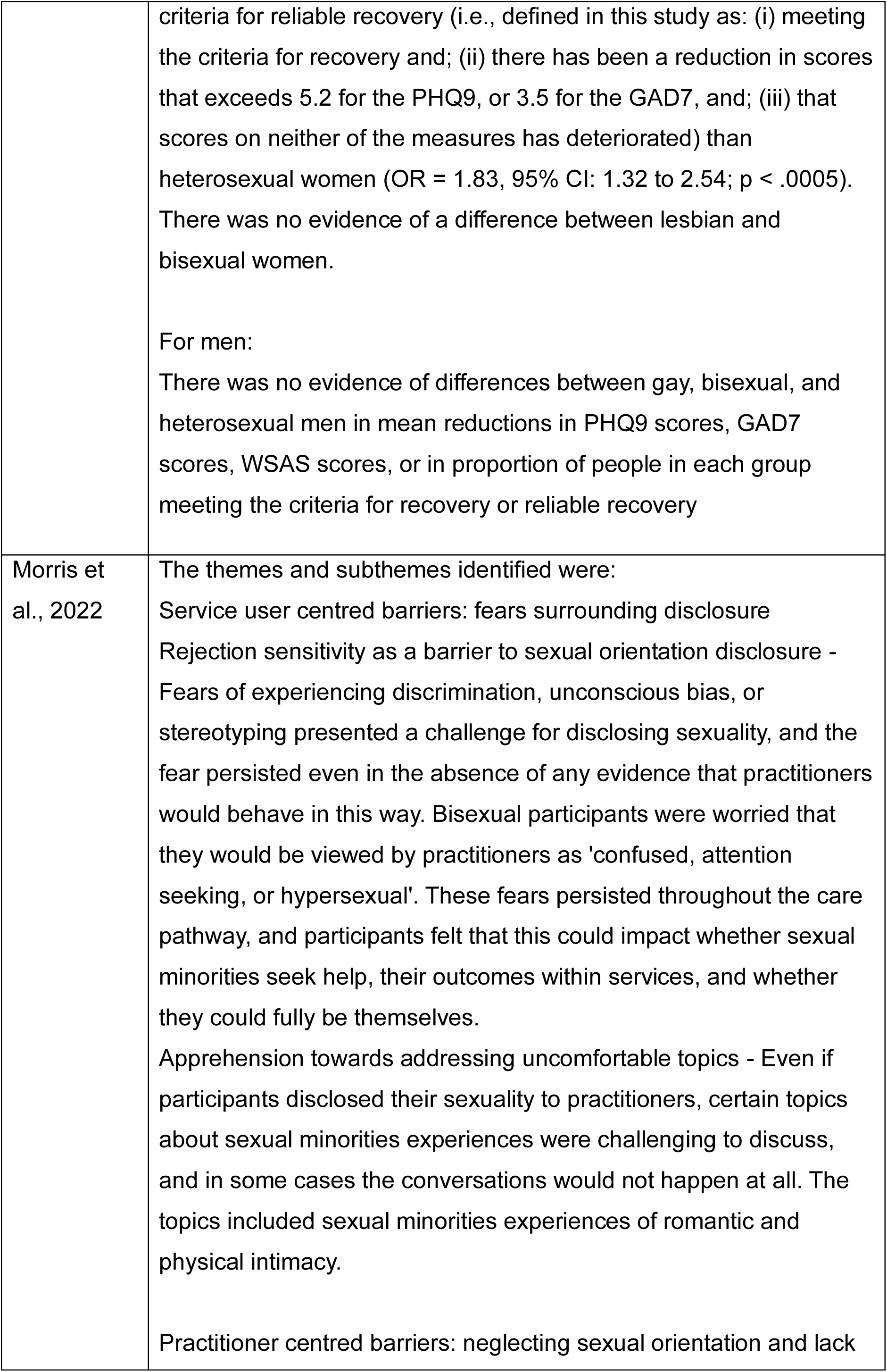
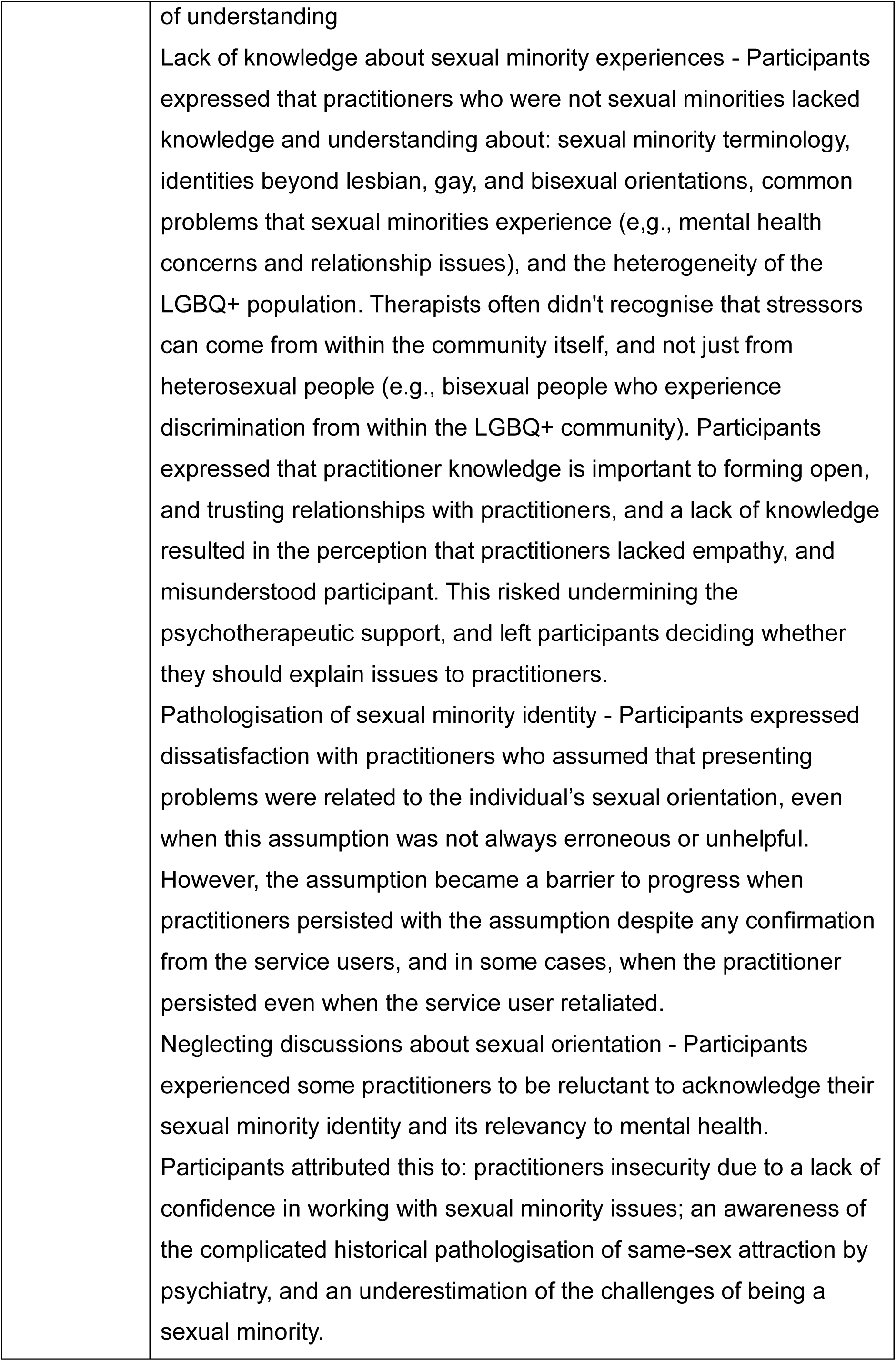
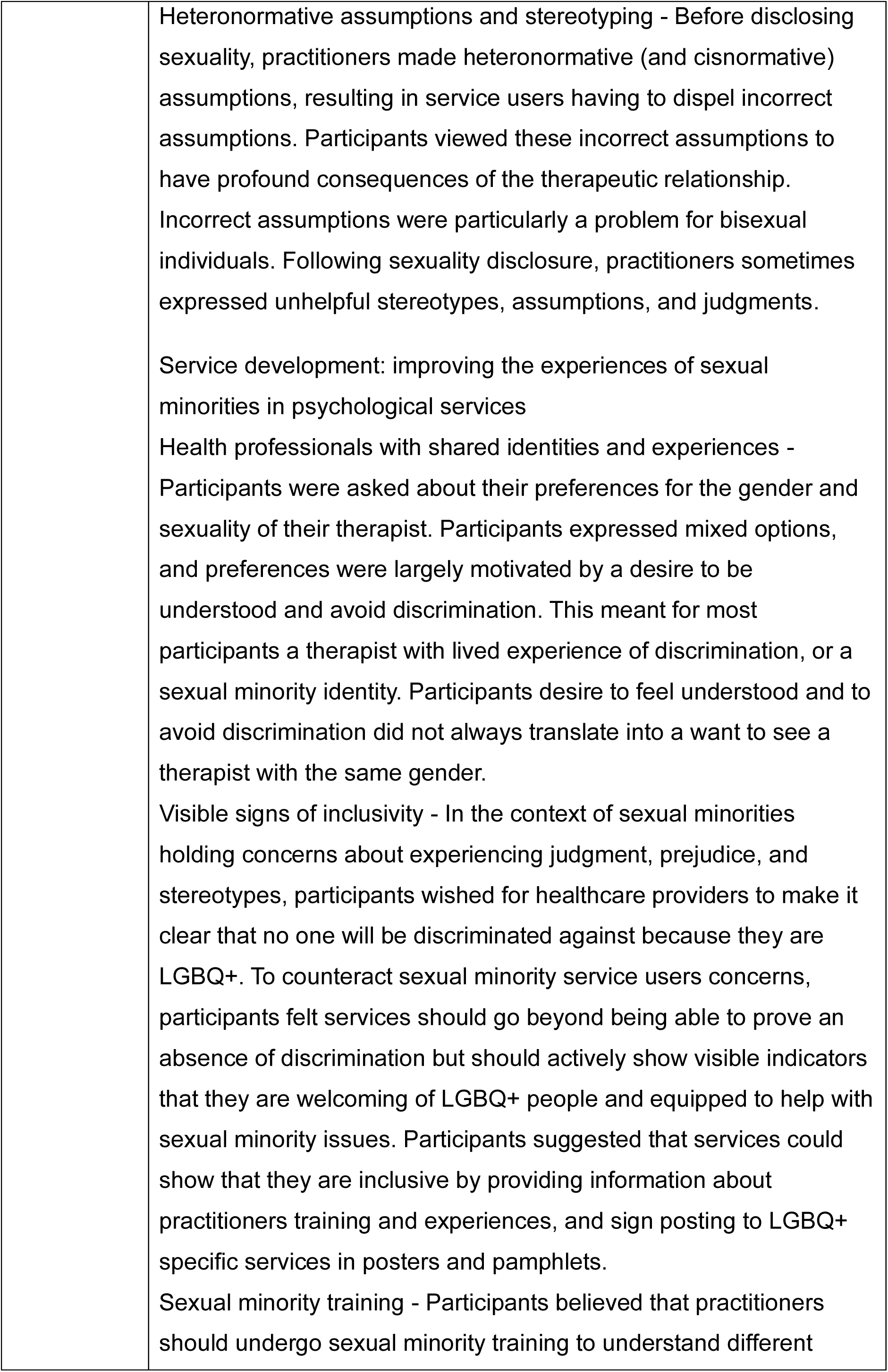
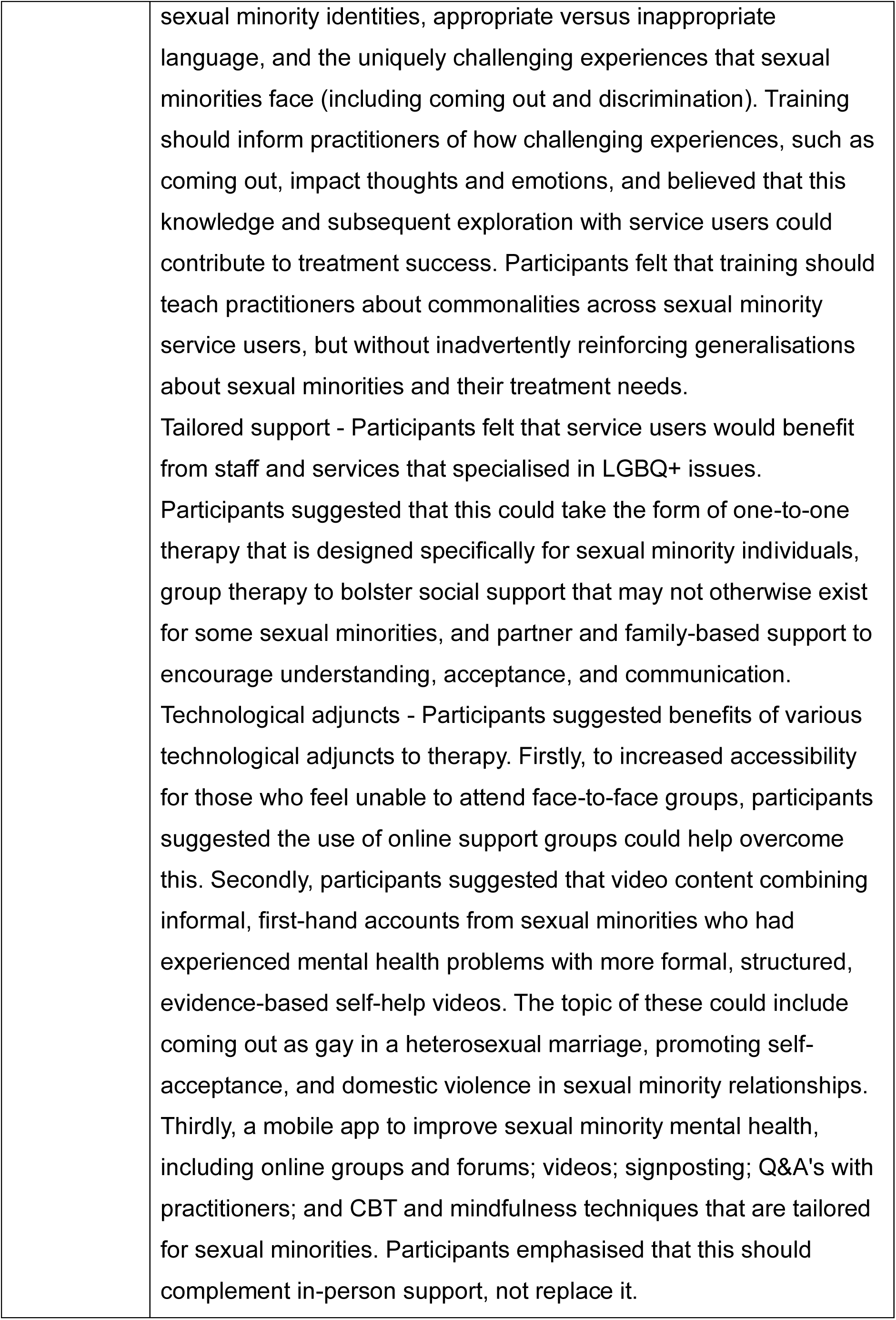
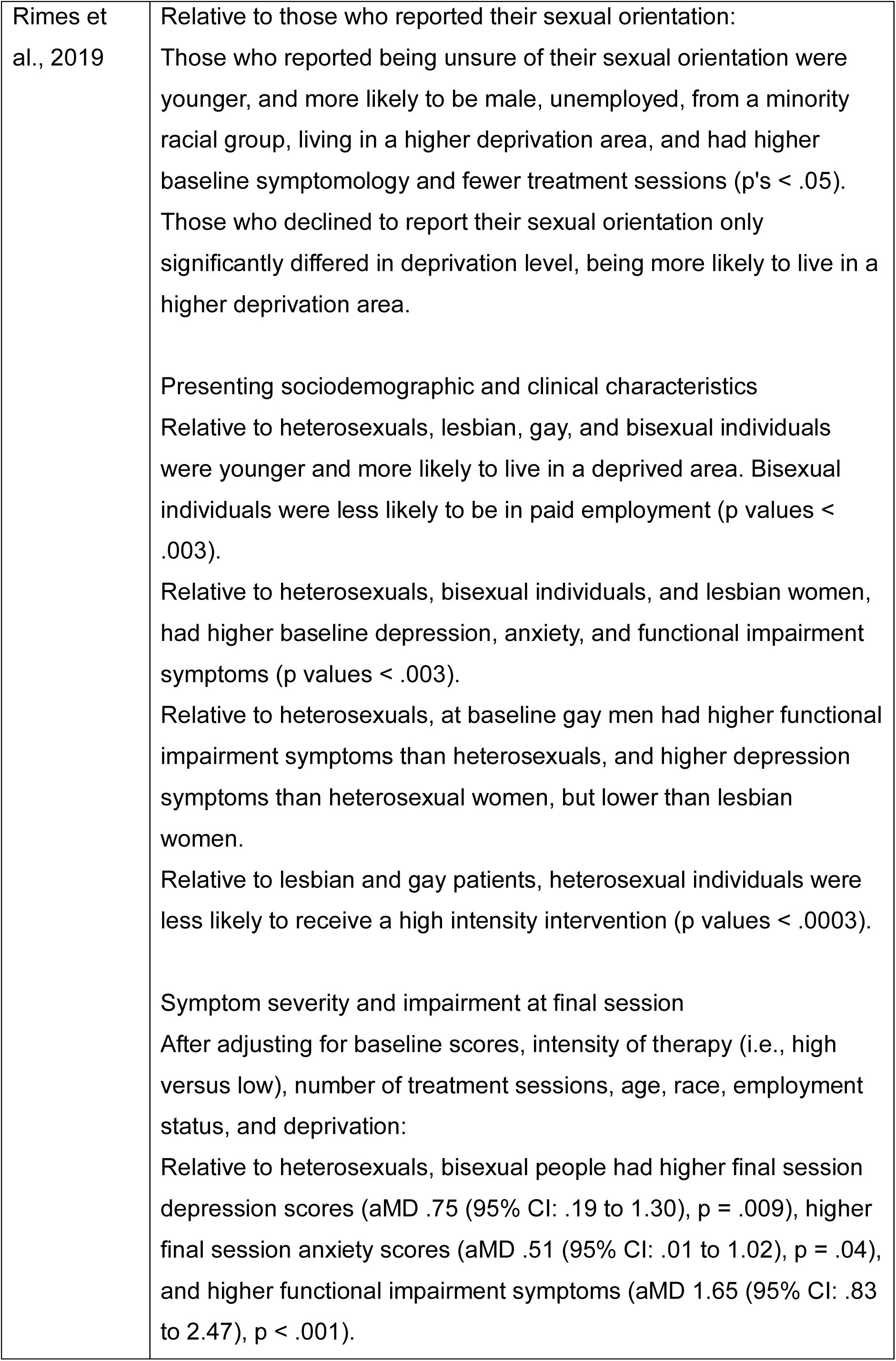
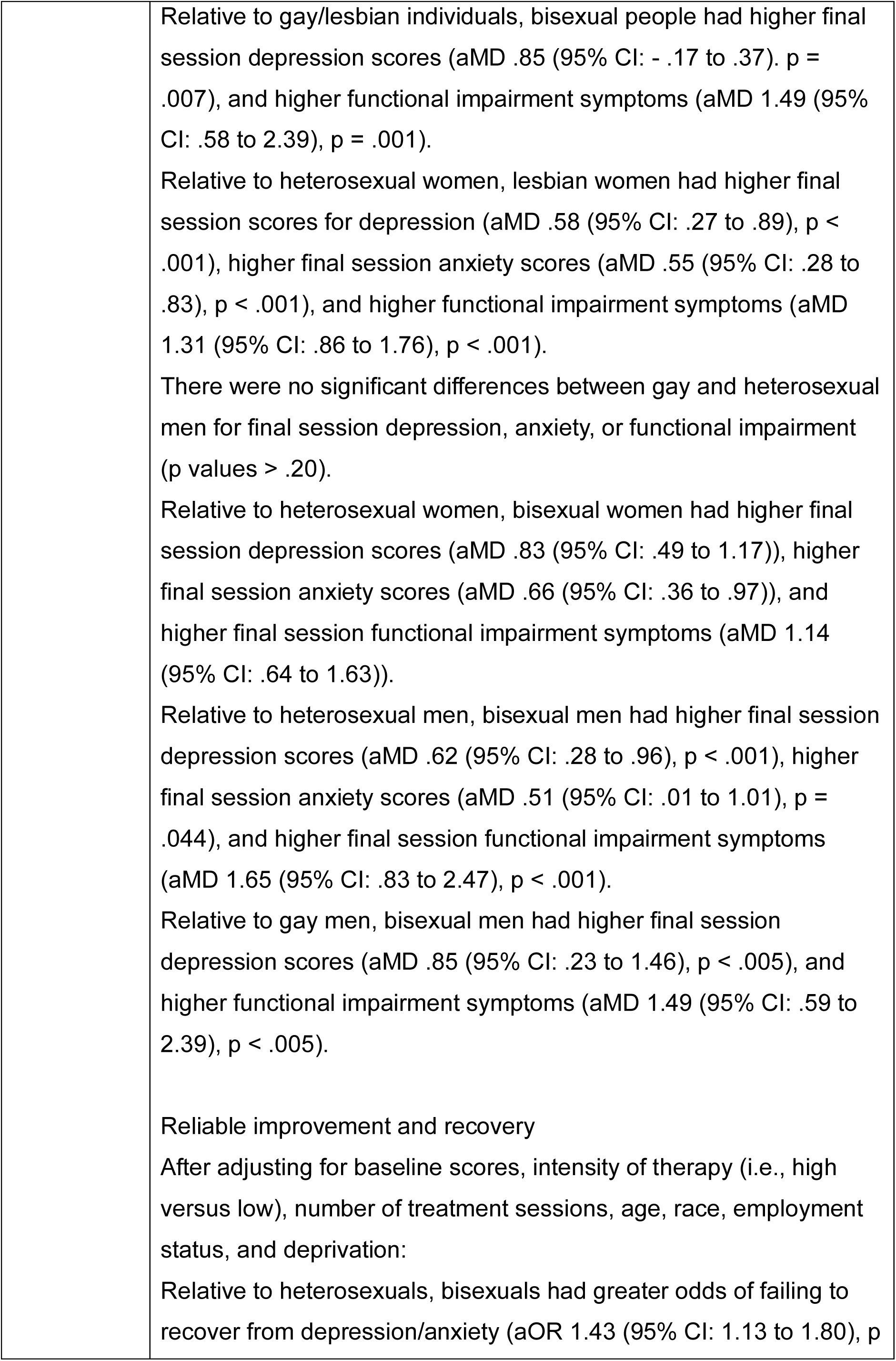
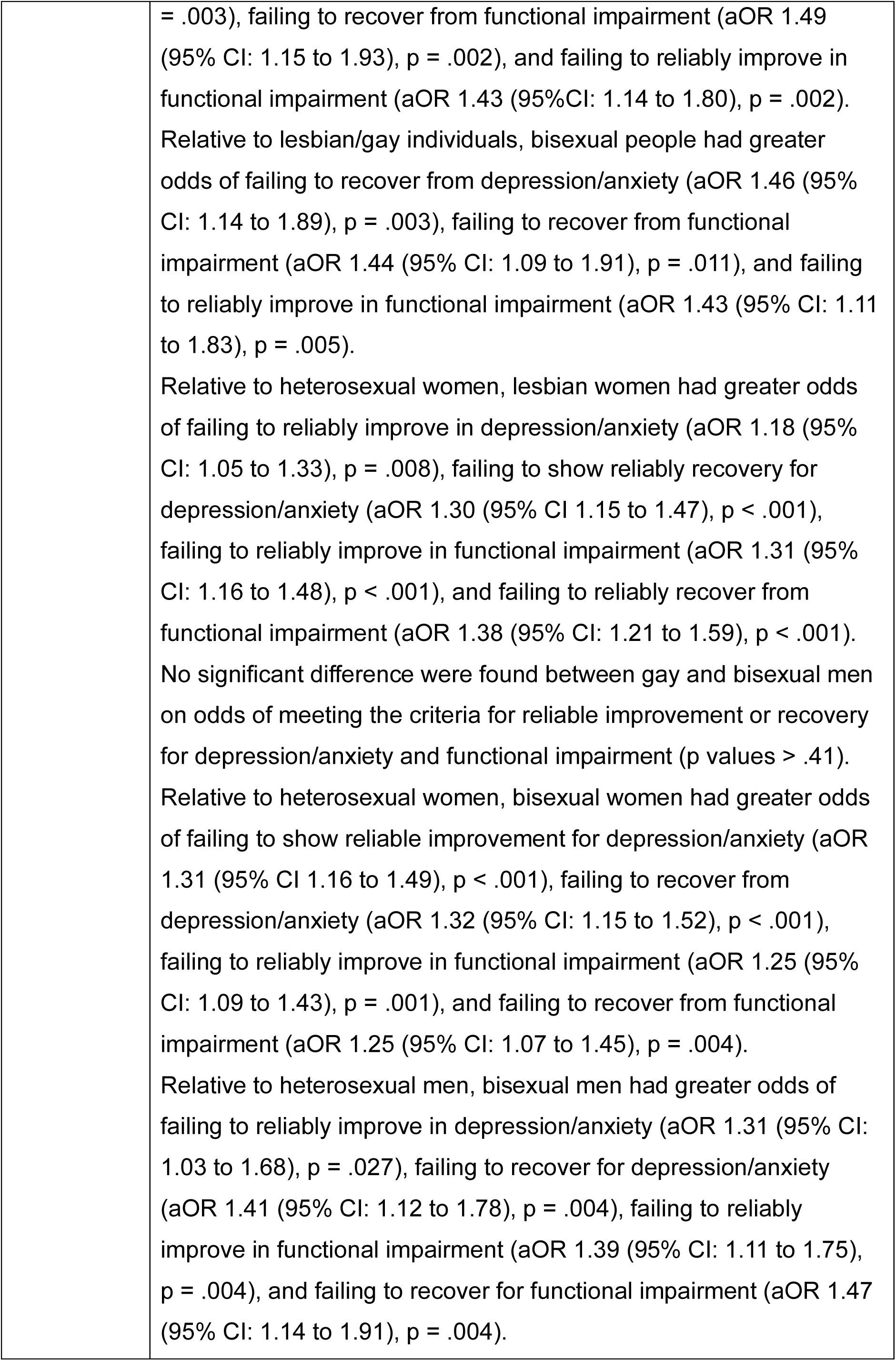
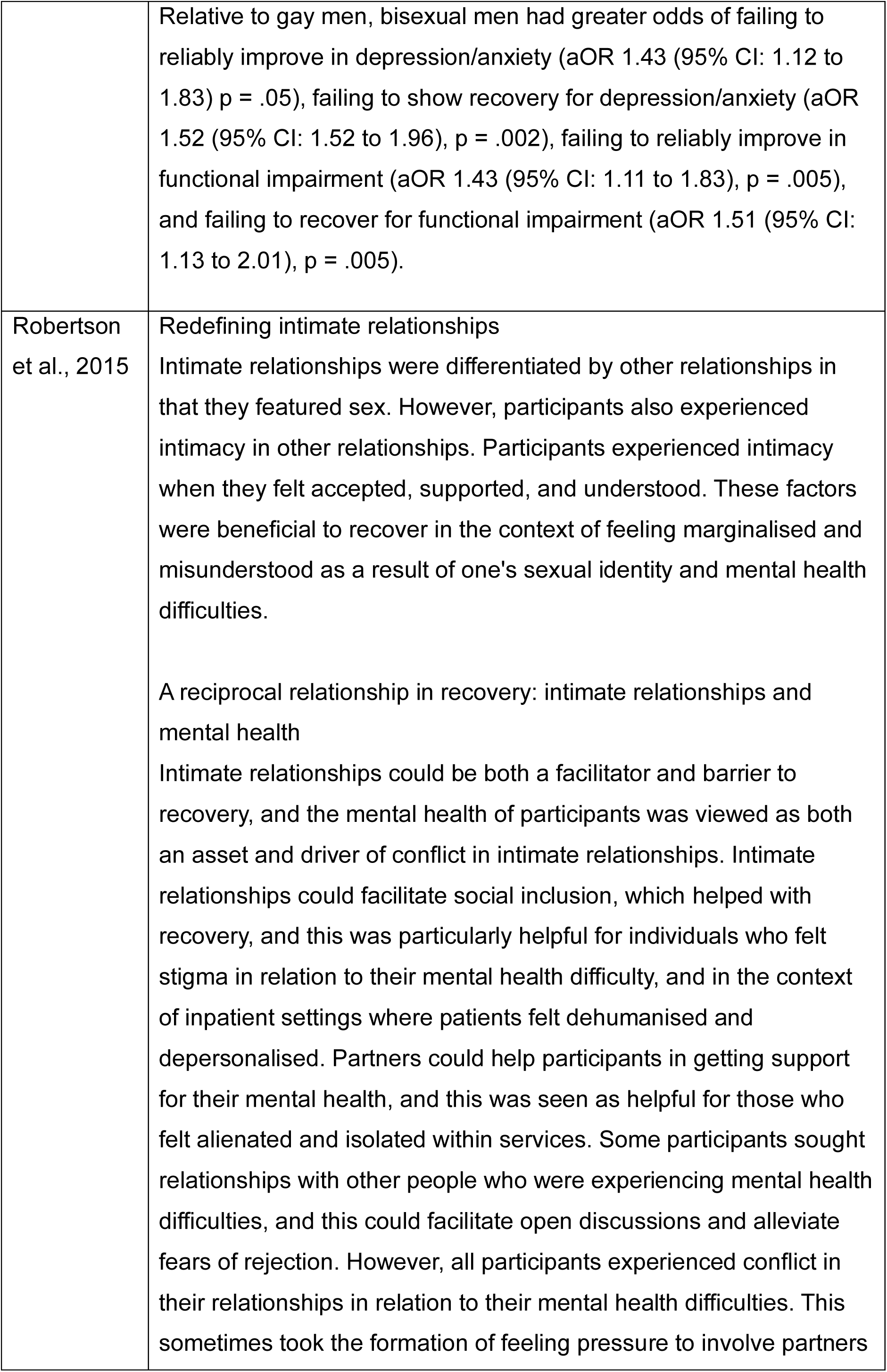
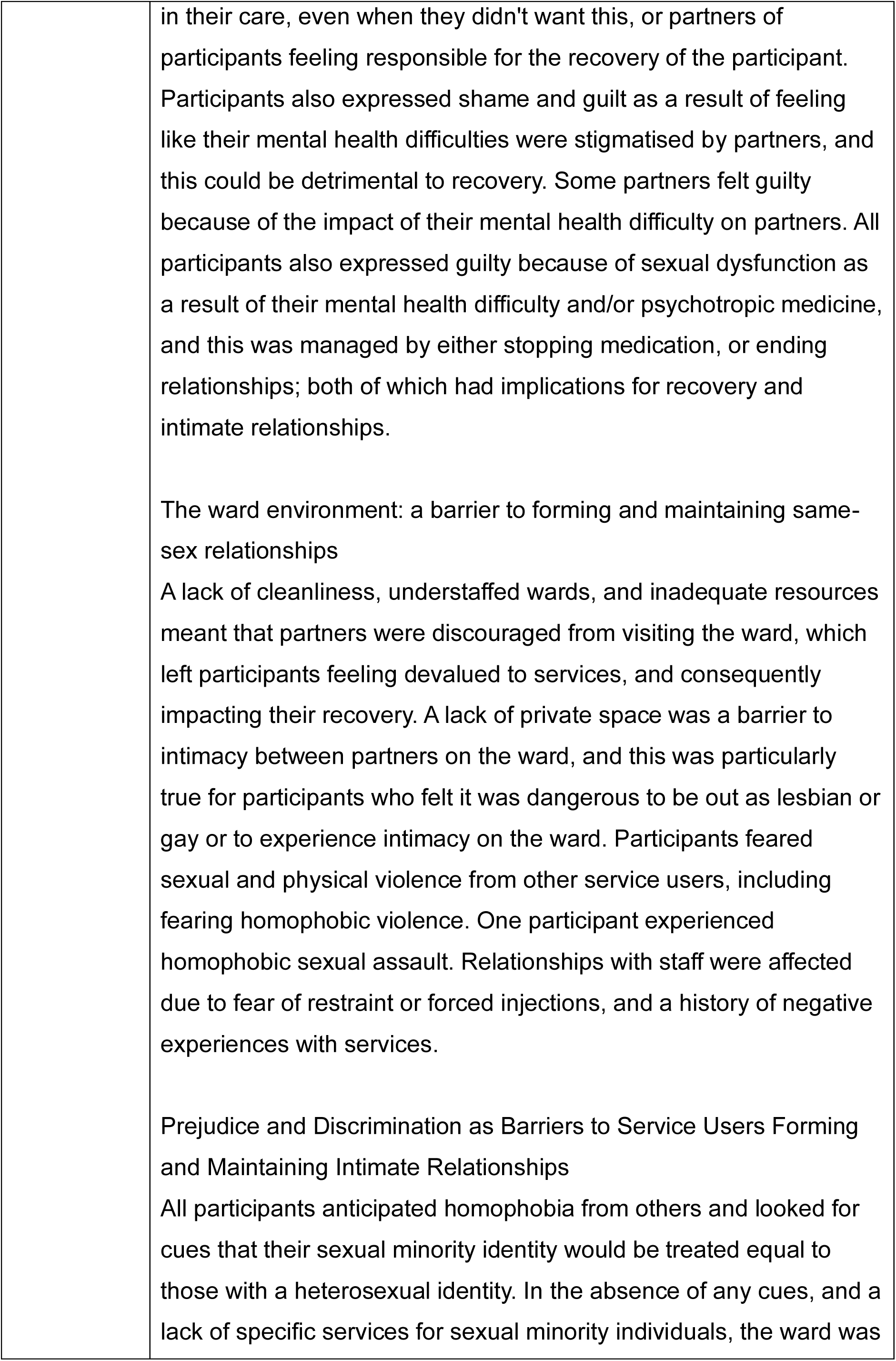
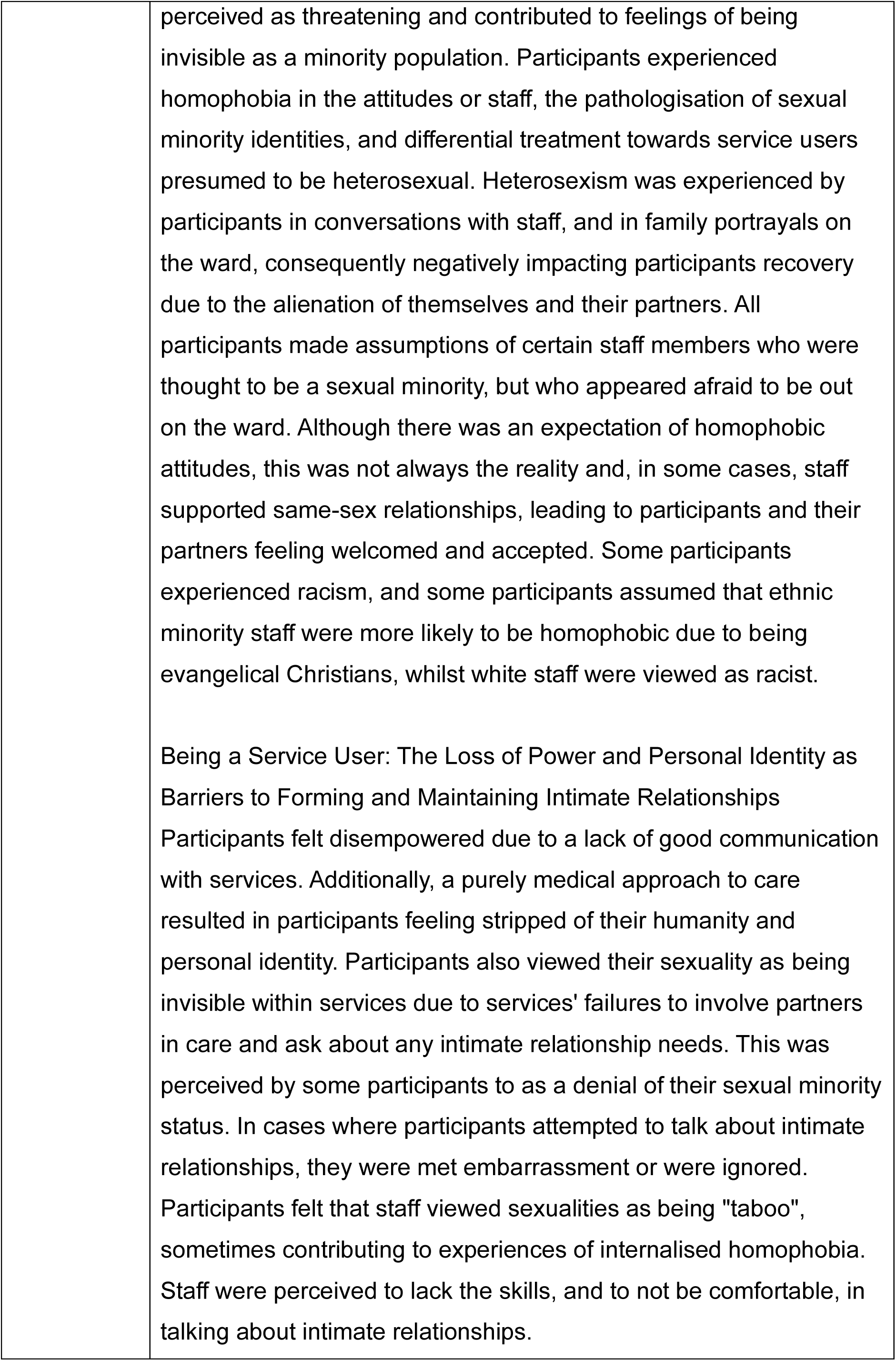
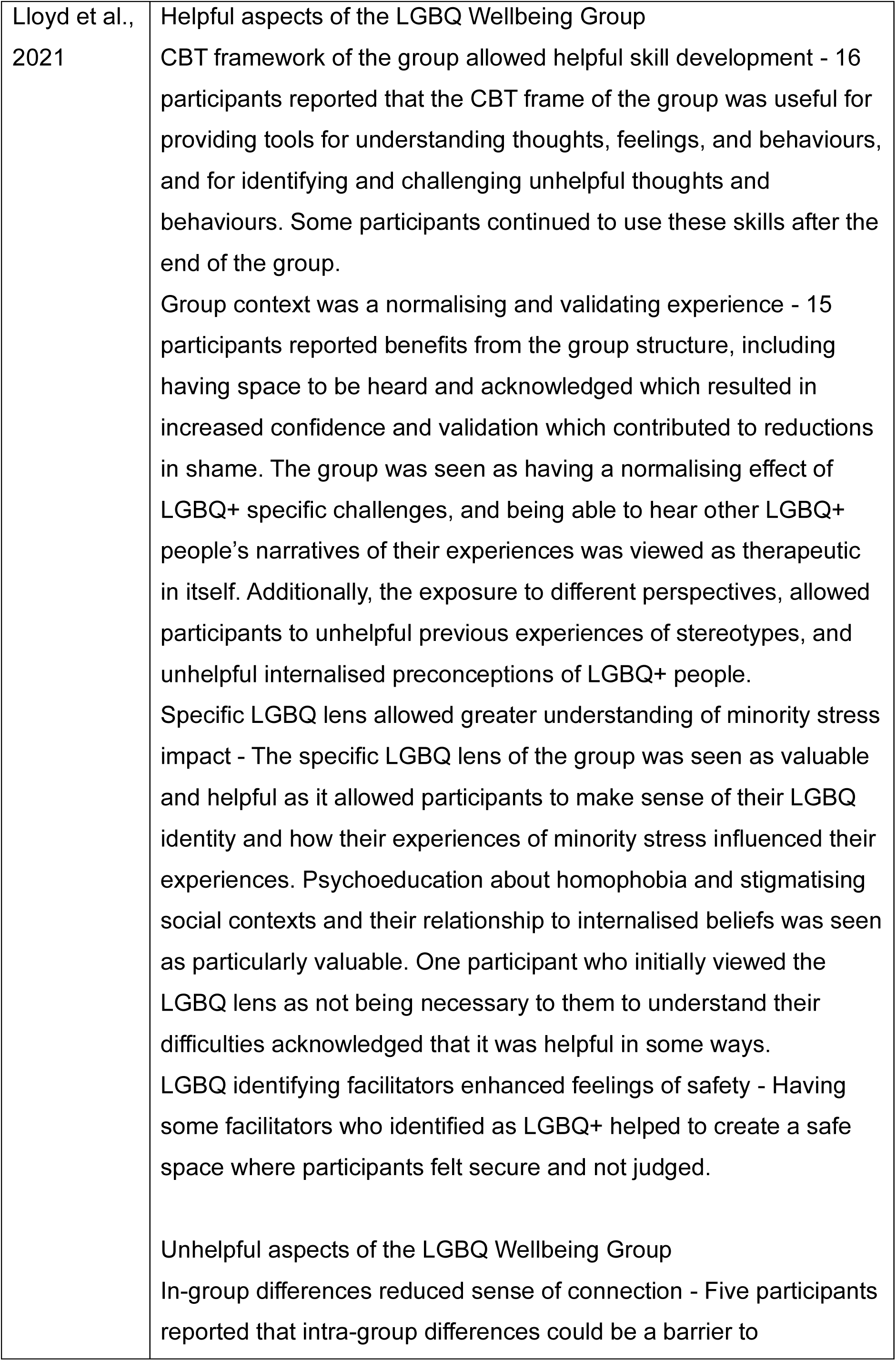
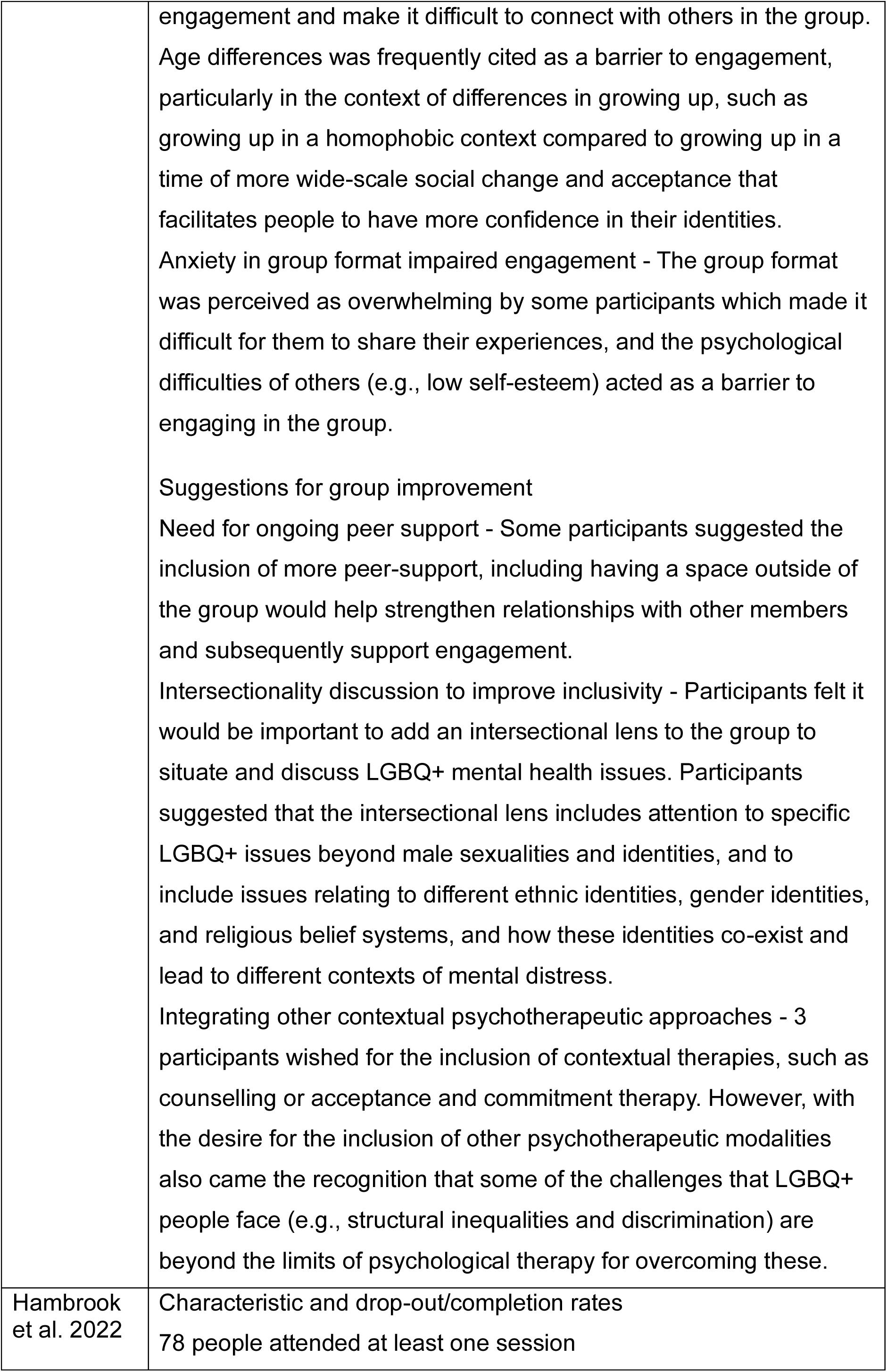
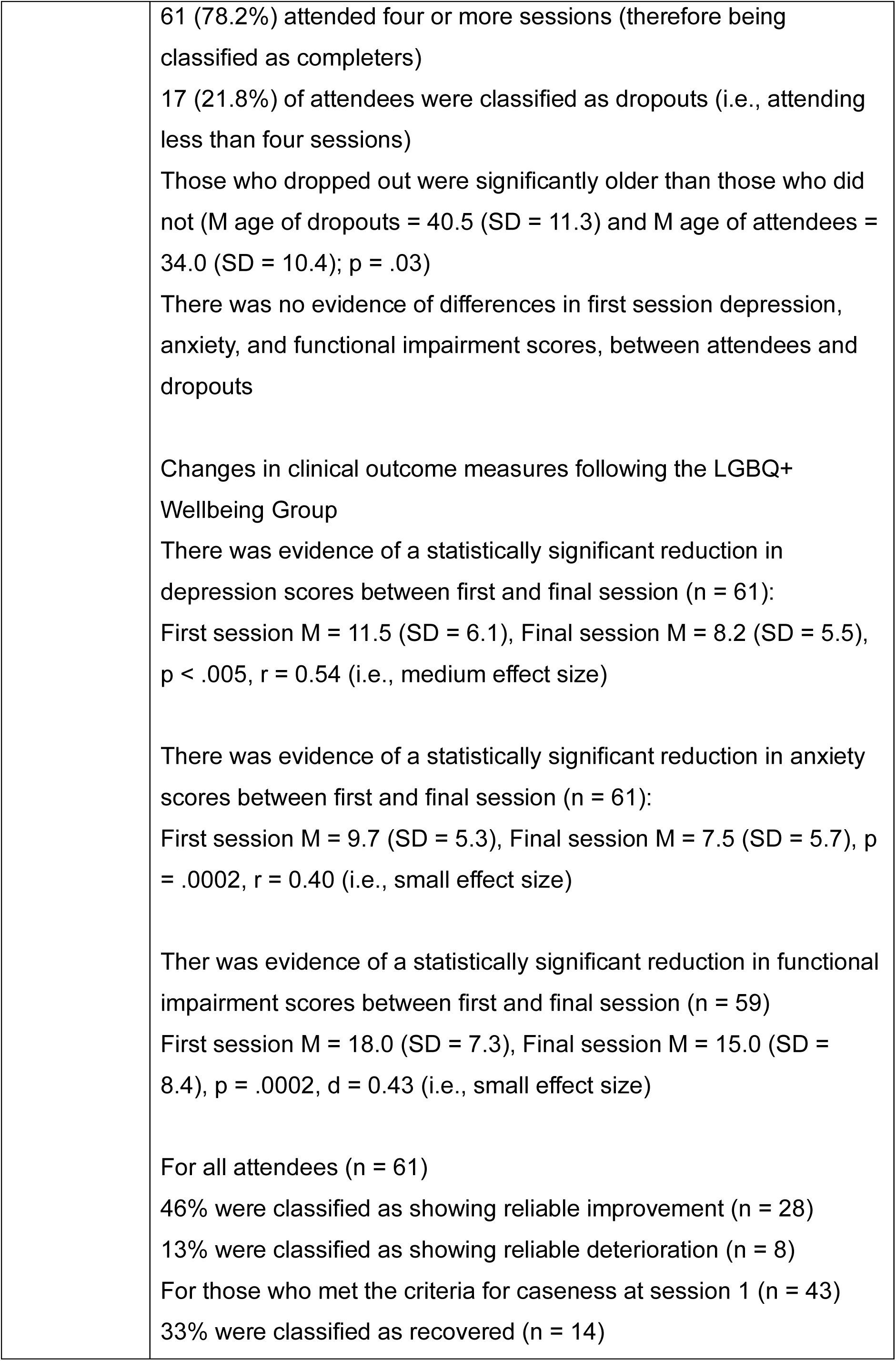
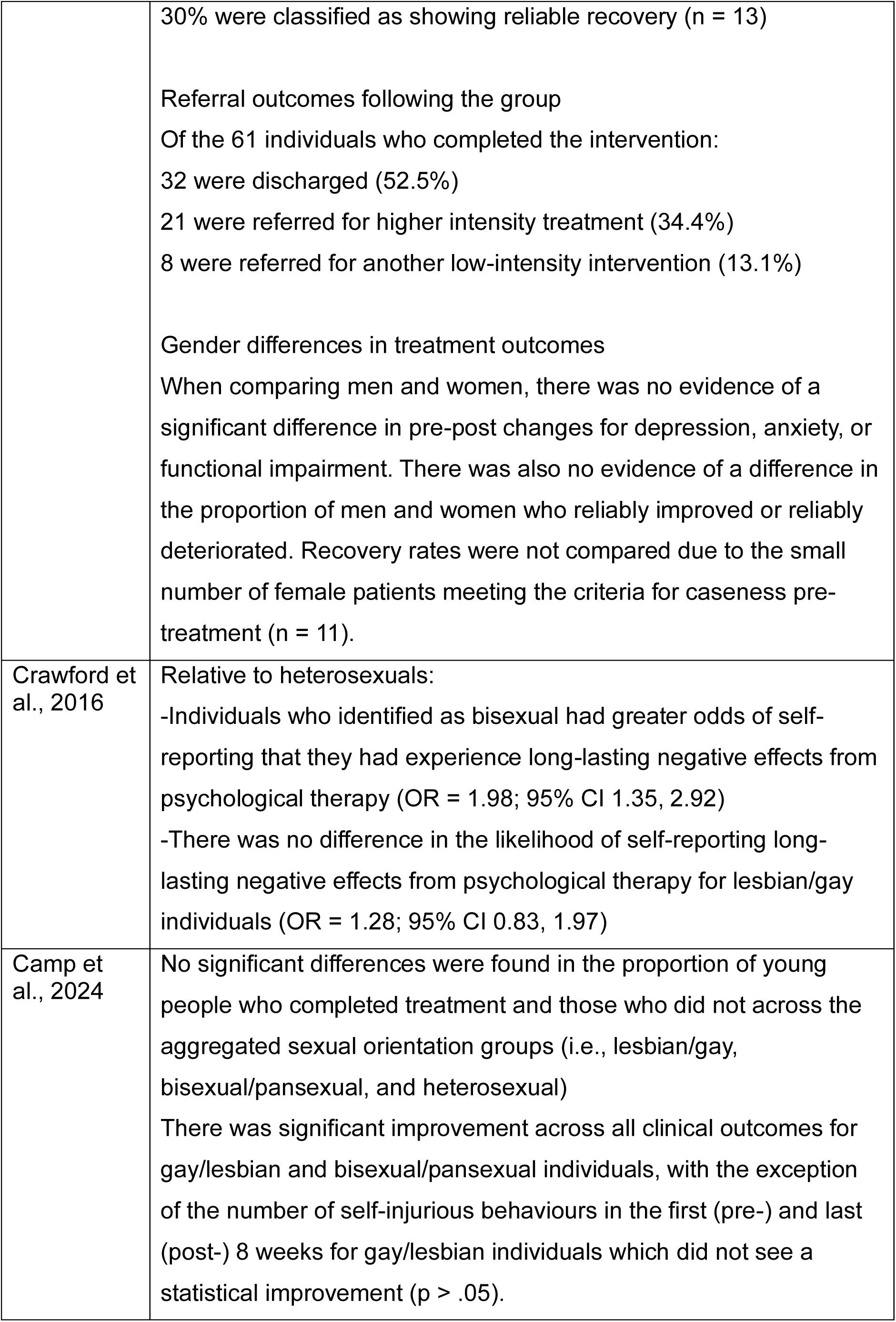
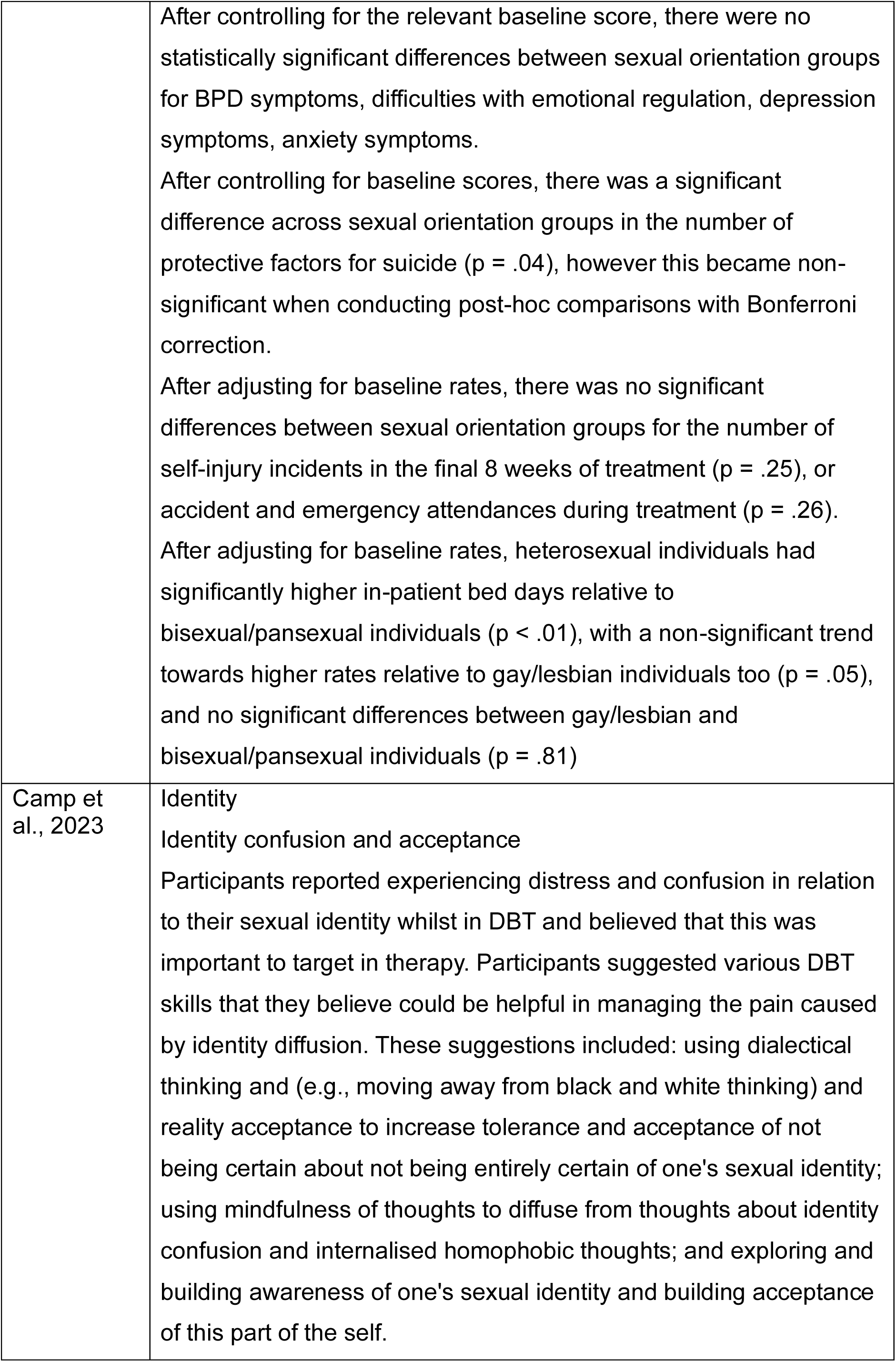
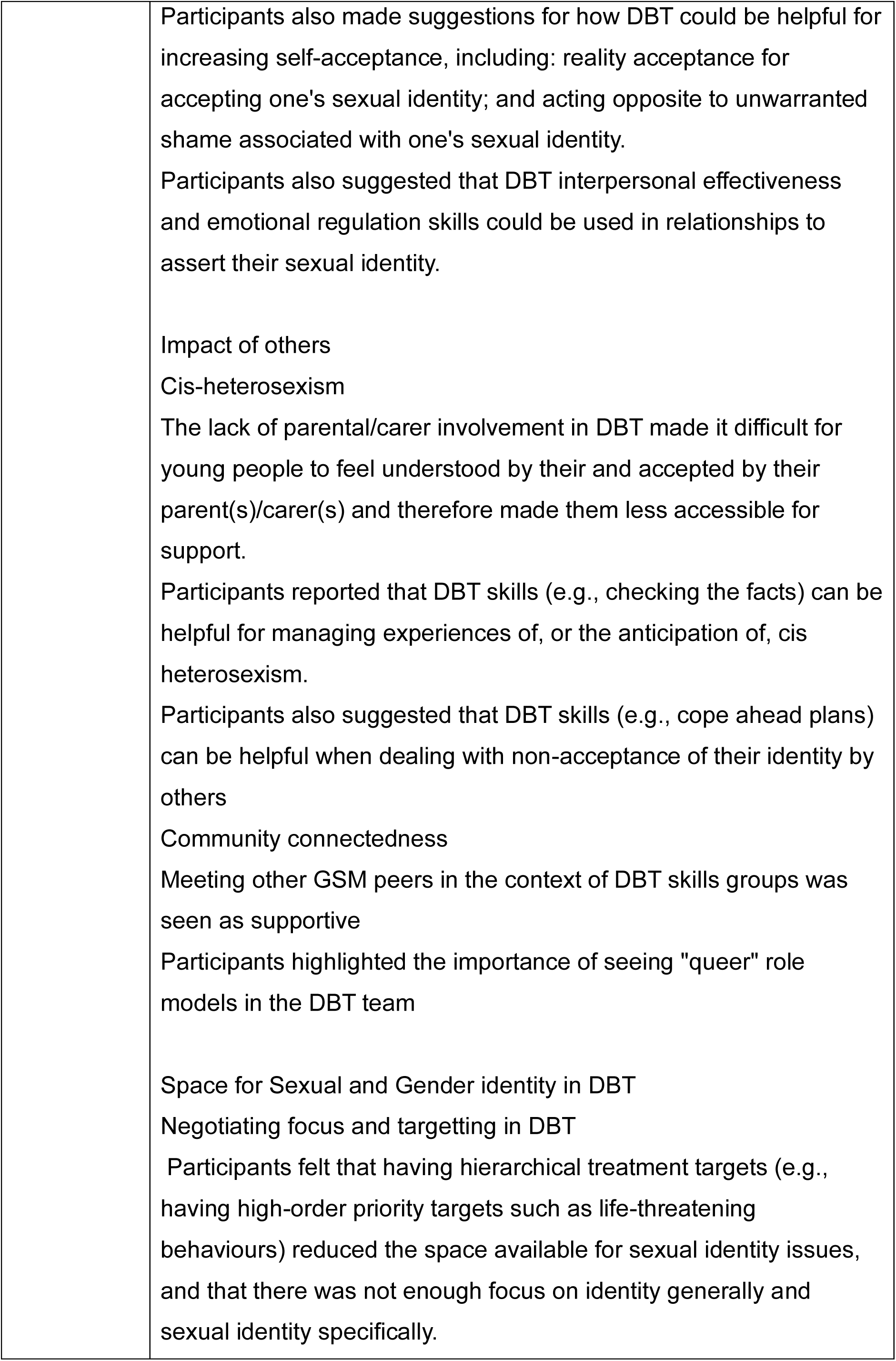
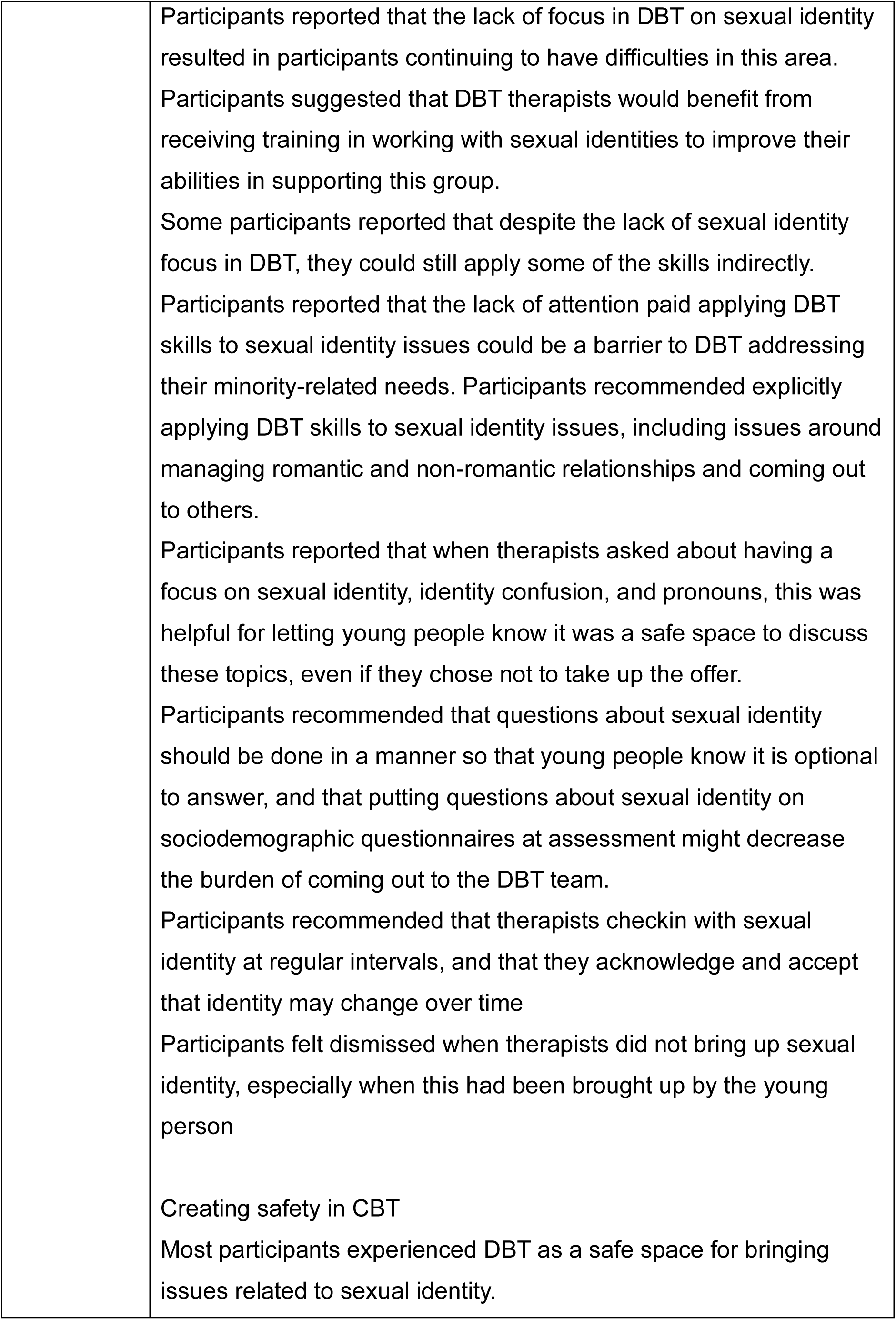
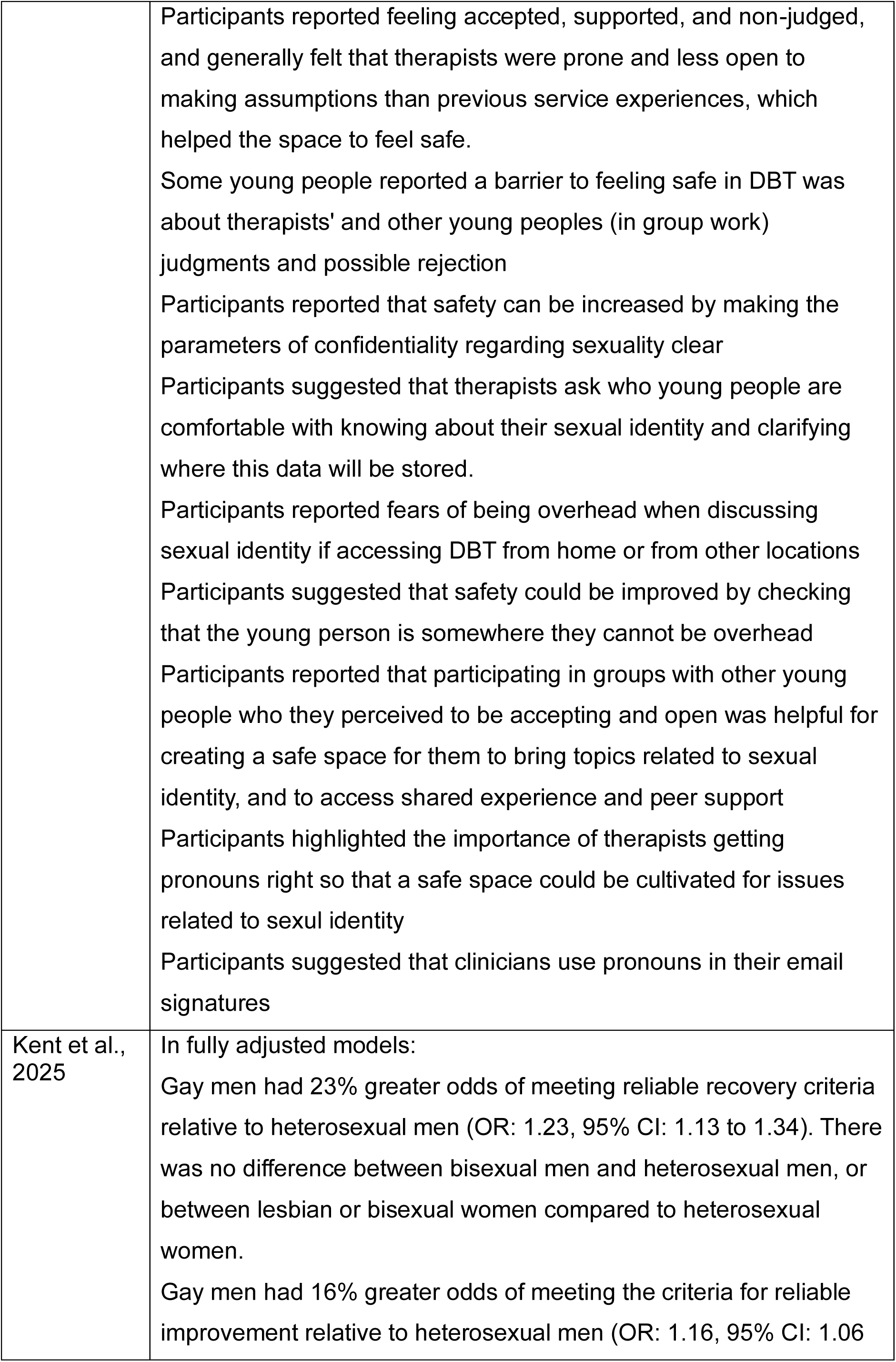
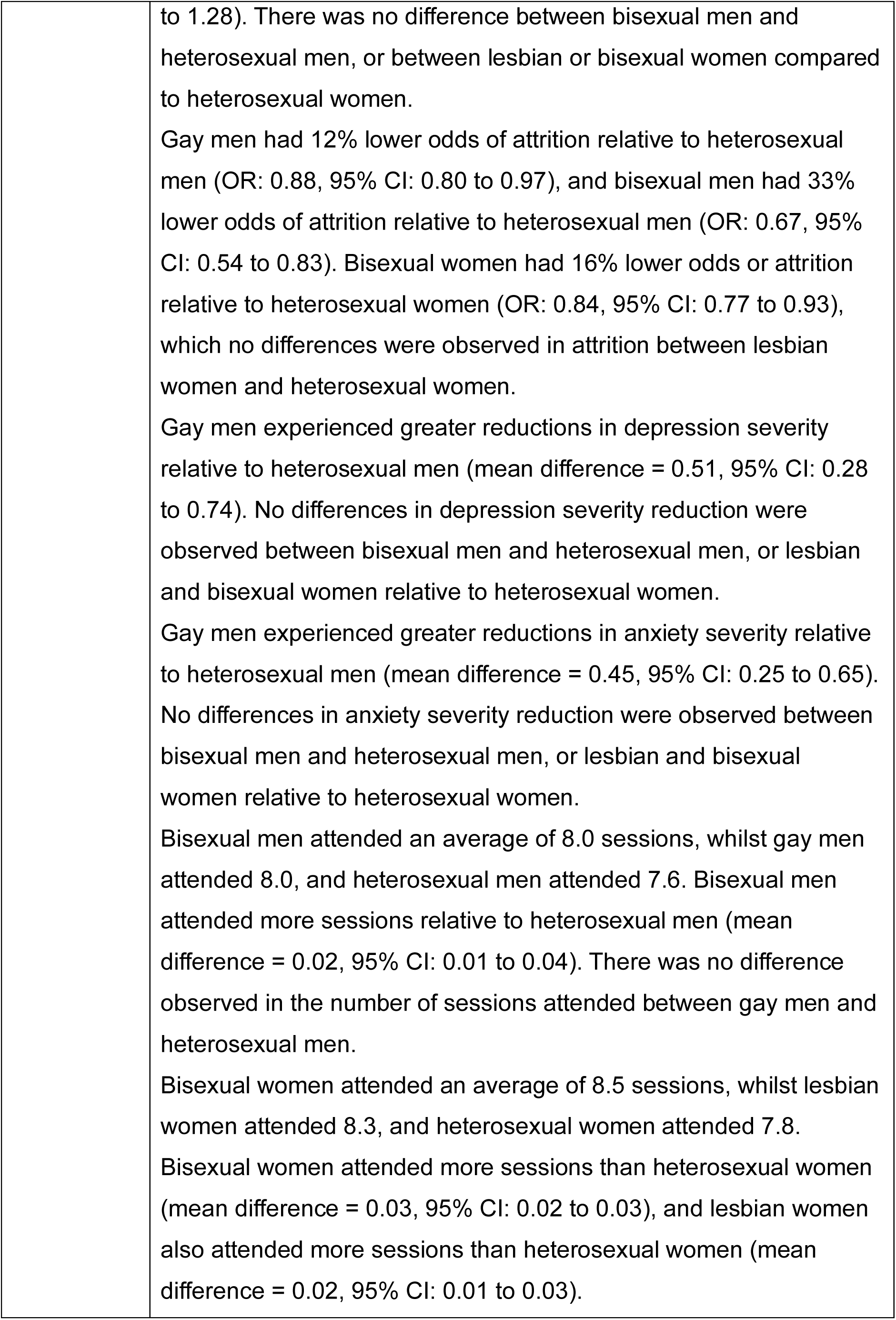

